# Ancestry-specific and multi-ancestry genome-wide association studies of restless legs syndrome

**DOI:** 10.64898/2026.04.28.26351960

**Authors:** Fulya Akçimen, Miranda Medeiros, Katie L.J. Cederberg, Marzieh Khani, Andy Roth, Mike A. Nalls, Sara Bandres-Ciga, Patrick A. Dion, Guy Rouleau, Emmanuel Mignot

## Abstract

Restless legs syndrome (RLS) is a common neurological disorder that disrupts sleep and quality of life, yet its genetics has been examined almost exclusively in individuals of European ancestry. We performed genome-wide association analyses of RLS in African (2,176 cases; 153,313 controls), Latin American (2,024 cases; 91,902 controls), and European (36,993 cases; 639,182 controls) ancestry groups, followed by a multi-ancestry meta-analysis. We leveraged biobank-based cohorts that established RLS diagnosis using validated clinical criteria, allowing for precise phenotypic characterization. We performed ancestry-specific association studies and post-GWAS approaches to prioritize candidate genes. Our analyses revealed ancestry differences in the genetics of RLS. At the *MEIS1* and *BTBD9* loci, lead variants showed lower allele frequencies and did not reach genome-wide significance in African ancestry. We identified ancestry-specific and shared risk loci, including novel associations near *GYPC/TEX51* and *PRIMA1* in African ancestry and *ISX* in Latin American ancestry. The European meta-analysis identified 11 additional loci and replicated 50 previously reported associations. The combined multi-ancestry analysis revealed ten new loci. This multi-ancestry study broadens the genetic understanding of RLS beyond European populations, revealing both shared and ancestry-specific contributors to disease risk. The absence or reduced frequency of key European RLS alleles in African ancestry individuals provides genetic insight into known epidemiological differences. Together, these findings lay the groundwork for mechanistic follow-up studies.

## Introduction

Restless legs syndrome (RLS) is characterized by an irresistible urge to move the legs, typically accompanied by unpleasant sensations. These sensations worsen in the evening and during periods of rest, such as lying in bed or sitting, and are relieved by movement. The condition impacts quality of life and impairs functioning in daily, occupational, and social domains^1,2^. RLS symptoms are common, with up to 10% of the European and North American population exhibiting some symptoms, and more severe in approximately 3% of the European and North American population^1^. The disorder is more common in women, and most affected individuals also exhibit periodic leg movements during sleep (PLMS), a condition that is characterized by involuntary leg movements that occur during sleep and can be detected through polysomnographic studies. Although several pharmacological options are available for RLS, long-term management can be challenging and often requires treatment adjustments due to side effects or loss of efficacy^3^.

Earlier family-based studies of RLS demonstrated significant familial aggregation^4^ and linkage analyses identified several candidate loci^5,6^. However, these loci were not replicated, and to date no causal variant has been described. Genome-wide association studies (GWASs) have uncovered a highly polygenic architecture for RLS and demonstrated shared genetic liability with PLMS^7^. Most genomic studies to date have been conducted in individuals of European ancestry^8,9,10,11,12,13^.

The most recent GWAS meta-analysis combined a clinically ascertained cohort (EU-RLS-GENE) with population-based datasets, most prominently those derived from survey-based resources such as 23andMe. This large-scale meta-analysis reported 193 independent variants in 161 associated autosomal loci. However, the majority of cases were derived from a single self-reported question regarding RLS status, rather than from a multi-item, criteria-based phenotyping approach, with clinically diagnosed RLS cases comprising a substantially smaller proportion of the total sample. As a result, differences in phenotyping approach across cohorts may have shaped the range of genetic associations observed^13^. Across European ancestry GWAS meta-analyses, *MEIS1* and *BTBD9* loci have consistently shown the strongest associations with RLS risk, together with numerous additional common variants with modest individual effects. Reported effect sizes for *MEIS1* vary across studies and study designs, with odds ratios of approximately 1.77-1.99 in European ancestry meta-analyses, and a larger effect (odds ratio = 3.71) observed in the clinically defined EU-RLS-GENE cohort. Similarly, *BTBD9* has been associated with modest effect sizes in population-based studies (odds ratios, approximately 1.26-1.35), with stronger effects observed in the EU-RLS-GENE cohort, corresponding to an odds ratio of approximately 1.48. Together, these findings indicate that numerous additional common variants confer modest effects, collectively underscoring the highly polygenic nature of RLS^9,13,14^.

In contrast, RLS has been far less studied in African and Latin American populations. A small number of prevalence-focused studies have suggested that RLS may be less frequently reported in some African populations, although estimates vary widely and are difficult to interpret given differences in study design, diagnostic criteria, healthcare access, and awareness of the disorder^15,16^. Importantly, the lower prevalence may reflect underdiagnosis, limited access to sleep medicine services, or broader healthcare inequities. In Latin American populations, prevalence data remain sparse and largely confined to small, region-specific studies^17,18^. RLS was examined as one of many traits in the Million Veteran Program GWAS, and *MEIS1* and *BTBD9* were replicated in the Latin American ancestry group, while no genome-wide significant associations were identified in individuals of African ancestry, in the context of comparatively low numbers of RLS cases in these ancestry groups^19^. Therefore, it remains unclear whether established RLS loci generalize across ancestries and whether additional ancestry-specific risk variants exist, which the present study was designed to address.

With the increasing availability of large-scale biobanks including individuals from diverse ancestries, it is now feasible to extend genetic studies beyond European populations. The primary objective of this study was to identify shared and ancestry-specific genetic risk loci for RLS using ancestry-stratified and multi-ancestry genome-wide association analyses. To this end, we performed ancestry-specific meta-analyses for African, Latin American, and European ancestry cohorts, as well as a large-scale multi-ancestry meta-analysis integrating results from each GWAS. Downstream analyses, including gene-based tests, tissue enrichment, and gene prioritization approaches, were performed to support biological interpretation.

## Results

### Ancestry-stratified genome-wide association studies across three ancestries

We first conducted GWAS in African, Latin American, and European ancestry cohorts from the All of Us program and the UK Biobank using individual[level, case-control data. Next, we conducted ancestry-stratified GWAS meta-analyses by including additional biobanks. For African and Latin American ancestry groups, we included data from the All of Us Research Program and the Million Veteran Program. For the European ancestry group, we included data from All of Us, UK Biobank, the Canadian Longitudinal Study on Aging, CARTaGENE, and the Million Veteran Program. GWAS in All of Us and UK Biobank were performed in the current study using individual-level case-control data, whereas previously published summary statistics were used for the remaining cohorts. African ancestry meta-analysis included 2,176 cases and 153,313 controls; Latin American meta-analysis included 2,024 cases and 91,902 controls, and European meta-analysis included 36,993 cases and 639,182 controls (<u>Table S1</u>, Figure 1). In the African and Latin American meta-analyses, only variants present in both the All of Us and Million Veteran Program cohorts were included, resulting in a total of 14,536,654 variants in the African meta-analysis and 8,254,303 variants in the Latin American meta-analysis. For the European meta-analysis, which comprised five datasets, we analyzed 9,350,470 variants that were present in at least three of the datasets. We defined genome-wide significance for these ancestry-stratified meta-analyses as *P*[<[5[×[10^-8^. Expected statistical power across ancestry groups is summarized in Table S3. Genomic inflation was minimal across ancestry groups after normalization to a standard sample size, with λ_1000_values of 1.03 for African, 1.01 for Latin American, and 1.005 for European meta-analyses (Table S4, Figure S2).

**Figure 1.**
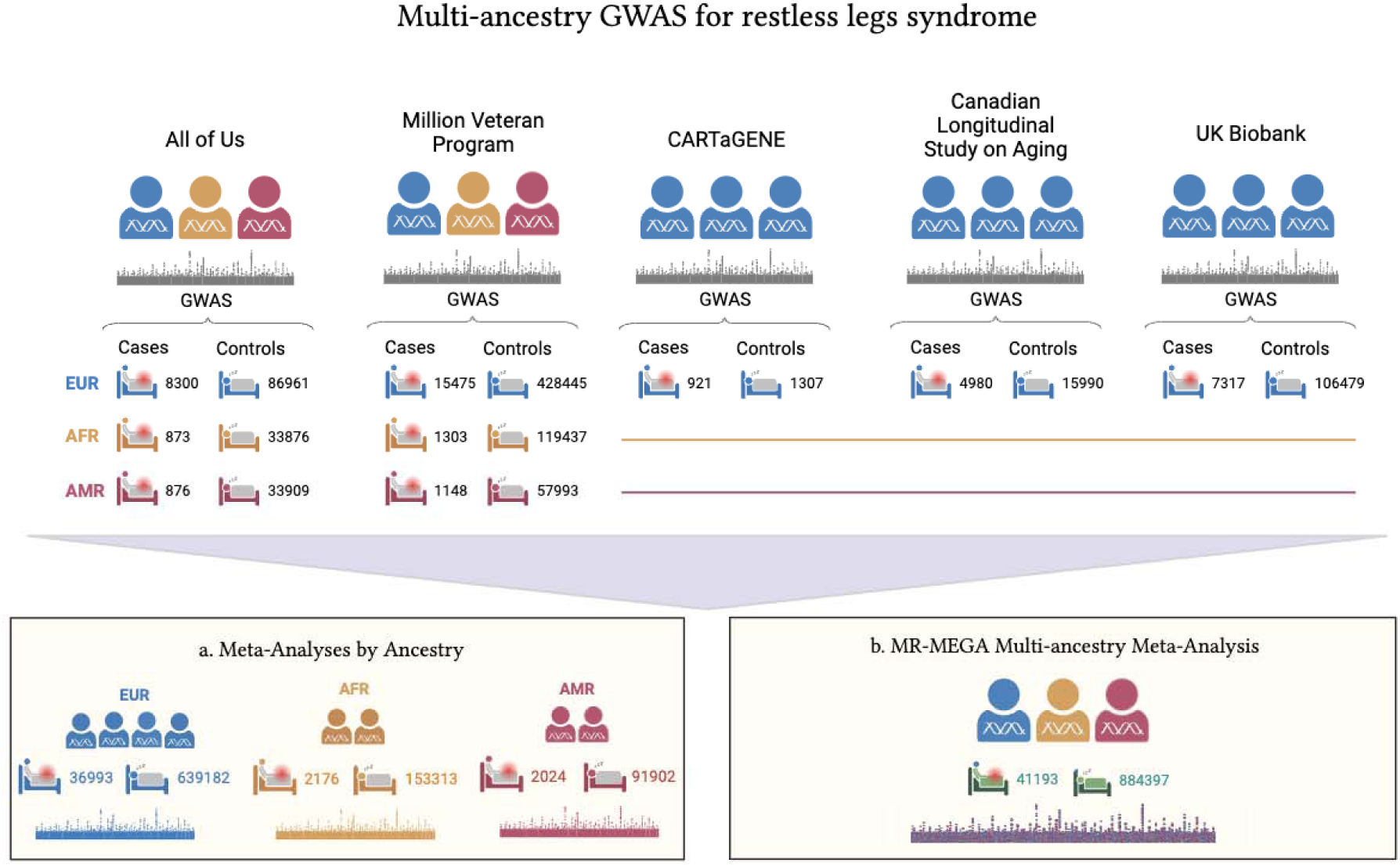
Overview of the multi-ancestry GWAS meta-analysis workflow for restless legs syndrome. Two meta-analysis approaches were used: a. ancestry-stratified meta-analyses were performed separately within African, Latin American, and European ancestry groups using METAL and b. A multi-ancestry meta-analysis of two African ancestry, two Latin American ancestry, and five European ancestry cohorts was performed using MR-MEGA.

In the Latin American ancestry meta-analysis, we replicated associations at the established RLS loci *MEIS1* (rs113851554-T) and *BTBD9* (rs9369062-C)^20^. For *MEIS1,* effect sizes were consistent across ancestries (AFR: OR=1.87, 95% CI=1.47-2.39, P=4.03 ×[10^-7^; AMR: OR=1.91, 95% CI=1.65-2.20, P=3.10[×[10^-18^; EUR: OR=1.95, 95% CI=1.90-2.01, P=1.25[×[10^-473^). For *BTBD9*, the genome-wide association was observed in AMR and EUR, while the effect in AFR did not reach significance (AFR: OR=0.92, 95% CI=0.82-1.02, P=0.12; AMR: OR=0.71, 95% CI=0.66-0.77, P=8.22[×[10^-20^, EUR: OR=0.77, 95% CI=0.76-0.78, P=5.66[×[10^-188^). We identified a novel locus near *ISX* (rs73166082-T), which reached genome-wide significance in AMR (OR=1.62, 95% CI=1.37-1.92, P=2.34[×[10^-8^ but not in AFR or EUR (AFR: OR=0.89, 95% CI=0.71-1.12, P=0.32; EUR: OR=0.99, 95% CI=0.95-1.04, P=0.76). In the African ancestry meta-analysis, we identified two genome-wide significant loci near the *GYPC/TEX51* (rs78603497-T; AFR: OR=1.77, 95% CI=1.44-2.16, P=3.15[×[10^-8^; AMR: OR=0.84, 95% CI=0.70-1.01, P=0.06; EUR: OR=1.01, 95% CI=0.98-1.04, P=0.63) and *PRIMA1* genes (rs10132474-G, AFR: OR=1.32, 95% CI=1.20-1.46, P=1.80 ×[10^-8^; AMR: OR=0.98, 95% CI=0.91-1.06, P=0.67; EUR: OR=1.002, 95% CI=0.98-1.02, P=0.80). These associations were not genome-wide significant in AMR or EUR ancestry (Table 1, Figure 2a-b, Figure S3).

**Figure 2.**
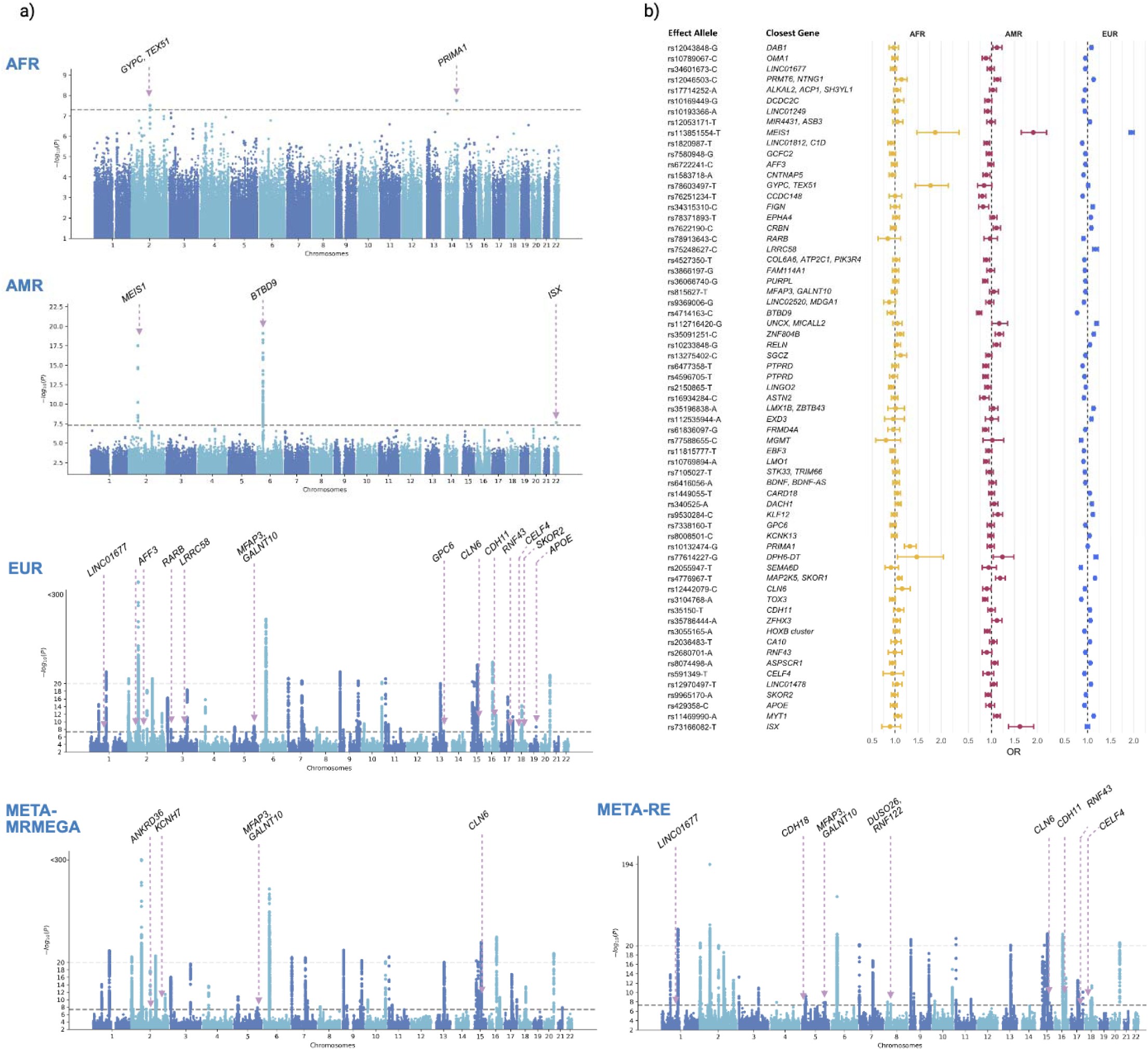
a. Manhattan plots for the three ancestry-stratified meta-analyses and the multi-ancestry meta regression results for restless legs syndrome. Only novel associations were labelled. b. Forest plot of lead RLS-associated variants across ancestries.

**Table 1.** Genome-wide significant associations in individual ancestry restless legs syndrome cohorts. Genome-wide significant associations in individual ancestry restless legs syndrome cohorts. Only novel variants were listed in European ancestry. Detailed summary statistics of genome-wide significant variants identified in ancestry-specific meta-analyses were listed in Table S5.

In gnomAD v4.1, the *MEIS1* and *BTBD9* risk alleles were five-fold and four-fold less frequent, respectively, in African and African American populations compared to European populations. For example, the *MEIS1* variant rs113851554-T had a frequency of 5.6% in the European population and 1% in the African and African American populations. Local ancestry inference among African and African American participants showed a frequency of 5.2% on European haplotypes but 0.02% on African haplotypes (<u>Figure S</u>4).

In the European ancestry meta-analysis, we identified 11 novel loci (near to *LINC01677*, *AFF3*, *RARB*, *LRRC58*, *MFAP3/GALNT10*, *GPC6*, *CLN6, CDH11*, *RNF43/TSPOAP1*, *CELF4*, and *SKOR2*) (Table 1, Figure 2, <u>Figure S3</u>) and replicated 50 previously reported GWAS loci^9,13^. In total, 83 independent variants were identified. Among these, rs34601673 (*LINC01677*) shows no linkage disequilibrium with the previously reported rs12044119 (R²=0.0011), rs815627 (*MFAP3/GALNT10*) shows no LD with rs10038916 (R²=0.0803), and rs12442079 (*CLN6*) shows no LD with rs868036 (R²=0.0804).

Ancestry-specific association statistics, frequencies, and effect sizes are reported in Table S5. Within the European ancestry meta-analysis, between-cohort heterogeneity was observed at a substantial number of known RLS loci, including *MEIS1* (I^2^=94.4), *BTBD9* (I^2^=84.7), *CCDC148* (I^2^=69.6), and *TOX3* (I^2^=68.3), likely due to cohort-specific factors such as differences in RLS case definition. Heterogeneity was also present at a subset of novel loci, such as *RARB* (I^2^=53.5), *AFF3* (I^2^=35.7), and *LRRC58* (I^2^=28.6), whereas other novel signals showed no heterogeneity (I^2^=0) (Table 1, Table S5). Three of the identified variants are protein-altering: p.Pro231Leu in *RNF43,* and p.Leu252Gln in *ASPSCR1*.

To determine whether previously reported loci are also observed in our independent dataset, we examined lead variants reported in prior RLS GWAS, including both the clinically diagnosed cohort and the meta-analysis incorporating 23andMe data. Of the 193 variants previously reported in the European meta-analysis, 45 showed significant association (p < 5 × 10^-8^) in our meta-analysis (Table S6). Among the remaining 148 variants that were not significant in our European meta-analysis, none reached genome-wide significance in the clinically ascertained EU-RLS-GENE consortium GWAS dataset (EU-RLS-GWAS) (Table S6)^13,14^. The most significant variants that failed to replicate were located in the *HLA* region: rs1059453-A (β = 0.116 ± 0.008, P = 1.71 ×10^-44^) and rs9269959-A (β = -0.120 ± 0.009, P = 1.20 × 10^-41^)^13^.

Notably, neither variant was significant in the previous EU-RLS-GWAS dataset^13,14^ (rs1059453-A, P = 0.053; rs9269959-A, P = 0.94) nor in our European meta-analysis (rs1059453-A, P = 0.62; rs9269959-A, P = 0.09) (Table S6).

In the ancestry-stratified meta-analysis, the *APOE* variant rs429358 (chr19:44908684:T>C), which defines the ε4 allele, was associated with reduced risk of RLS in individuals of European ancestry. We next examined the association between *APOE* genotypes and risk of RLS across ancestry groups (Table S7). In the UK Biobank European cohort, an increasing ε4 allelic dosage was associated with reduced RLS risk and a similar direction of effect was observed in the All of Us European cohort. Because this signal was largely driven by the UK Biobank, where RLS status was derived from an optional questionnaire, we considered the possibility of participation bias. Participation in UK Biobank optional components is known to be non-random and influenced by genetic factors. Recent work has demonstrated that multiple loci, including variants associated with intelligence and Alzheimer’s disease, are strongly associated with participation in optional surveys, highlighting a broader, polygenic source of selection bias rather than a trait-specific effect^21^. To assess whether the *APOE* association could reflect this broader participation bias, we performed a GWAS of participation in the UK Biobank RLS diagnostic survey. The rs429358 variant was strongly associated with questionnaire participation (OR = 0.89 [0.88-0.90], P = 6.52 × 10^-75^). Other identified RLS risk variants did not show associations with participation (Table S8). Age-stratified analyses showed a stronger inverse association at *APOE* variant in older participants (>=60 years: OR=0.84 [0.77-0.92]; P = 1.6 × 10^-4^ than in younger participants (<60 years: OR=0.92 [0.87-0.97]; P = 0.0043). A full list of variants reaching genome-wide significance is provided in Table S8.

We examined whether 83 lead variants were associated with variation in age at onset or symptom frequency in cohorts with available individual-level data. Age at onset analyses were performed among RLS cases only in the All of Us cohort. We observed a significant association for the *MEIS1* lead variant rs113851554, with the risk allele associated with a later age at onset (β = 1.54 years per allele, SE = 0.36; P = 2.03 × 10^-5^), exceeding the Bonferroni-corrected significance threshold (P < 0.0006, α = 0.05 / 83). This effect corresponds to an average increase of approximately 1.5 years in age at onset per risk allele (Figure S5). No other lead variants showed significant associations with age at onset across ancestry groups (Table S9). In analyses of symptom frequency among RLS cases in the UK Biobank, modeled as an ordinal outcome, we did not observe significant associations between lead variants and symptom frequency after correction for multiple testing (Table S10).

### Multi-ancestry GWAS meta-analysis

We aggregated summary statistics from two African ancestry, two Latin American ancestry, and five European ancestry cohorts using a multi-ancestry meta-regression (MR-MEGA)^22^, which incorporates study-level ancestry principal components derived from allele frequency differences to account for variation in effect sizes across ancestries. We also performed an inverse-variance weighted random-effects meta-analysis in PLINK v2^23^. Combining results from the random-effects model and MR-MEGA, we identified 10 novel RLS risk loci (Table 2) and confirmed 53 hits in previously reported loci from European ancestry GWASs (Table S12, Table S13). Notably, *LINC01677*, *MFAP3/GALNT10*, *CLN6, CDH11*, *RNF43*, *CELF4*, and *SKOR2* overlapped with novel signals from our European ancestry GWAS, whereas *ANKRD3, KCNH7,* and *DUSO26/RNF122* loci reached genome-wide significance exclusively in the multi-ancestry GWAS (Figure 2a-b, Table S4, Table S12, Table S13).

**Table 2.** Novel loci identified by MR-MEGA or random effect multi-ancestry meta-analysis for restless legs syndrome. Lead variants reaching genome-wide significance in either MR-MEGA or the PLINK random-effects (RE) meta-analysis are shown. Each method identified a different lead variant at the *MFAP3-GALNT10* and *CLN6* loci; both are listed. EA:effect allele; P (MR-MEGA): MR-MEGA association P value; P (RE):Random effect meta-analysis P value; I^2^ (RE):between-study heterogeneity (%)

### Fine mapping of genetic signals

We performed cross-ancestry fine-mapping using SuSiEx to identify likely causal variants underlying genome-wide significant loci (Table 3). SuSiEx integrates linkage disequilibrium (LD) patterns and effect size estimates across ancestries to improve fine-mapping resolution and identify shared causal variants^24^. Across all tested regions, we identified 14 credible causal variants with a posterior inclusion probability (PIP) > 0.8, of which ten loci contained a single variant.

**Table 3.** Cross-ancestry fine-mapping identified credible causal variants identified for risk signals of restless legs syndrome. Variants with a posterior inclusion probability[>[0.8 are reported.

For most loci, the lead variants identified in the GWAS were also fine-mapped as the most credible causal signals. For *MEIS1*, *LINC01812/C1D* and *BTBD9*, the fine mapped variants are in strong LD with the top association hits. At the *MEIS1* locus, the fine-mapped variant rs11679120 is highly correlated with the lead GWAS variant rs113851554 (D′ = 0.98, r^2^ = 0.80). At the *MEIS1* locus, the 95% credible set contained a single variant, rs11679120 (credible set size = 1; PIP = 1). Regional association plots (Figure S3c), displaying the linkage disequilibrium structure relative to the selected reference variant, show that rs113851554 forms the primary association peak, whereas rs11679120 lies within a nearby LD cluster of correlated variants in European and Latin American datasets. Conditional regression analysis in the European dataset showed that rs11679120 loses significance after conditioning on rs113851554 (OR = 0.87, P = 0.027), while rs113851554 remains strongly associated after conditioning on rs11679120 (OR = 2.04, P = 4.1 × 10^-38^), indicating that both variants represent the same underlying association signal. Similarly, at the *BTBD9* locus, the fine-mapping prioritized rs9369062. This variant was in high LD with the European lead variant rs4714163 (D′ = 1, r^2^ = 0.98), which was also the lead variant in the Latin American GWAS. Because these variants are essentially indistinguishable statistically, conditional regression could not be reliably estimated, consistent with a shared causal signal.

### Gene-based association and tissue-specific expression enrichment

After correction for multiple testing, 49 genes were significantly associated with RLS in the European cohort, whereas only *BTBD9* reached significance in the Latin American cohort. No significant gene-level associations were observed in the African ancestry cohort. In the random-effects meta-analysis dataset, 26 genes were significantly associated with RLS (Table S14).

We examined tissue specific expression patterns of RLS-associated genes using MAGMA, based on GTEx v8 transcriptomic data across 53 tissue types. In the European GWAS, we observed a significant enrichment for genes predominantly expressed in the cerebellum and cerebellar hemisphere. We did not observe significant enrichment in the African or Latin American ancestry analyses, nor in the multi-ancestry meta-analysis (Figure S6).

### Colocalization, transcriptome-wide association and summary-based Mendelian randomization analysis nominate eight putative genes

Across all genome-wide significant loci in the current study, the six colocalization and transcriptome-integration approaches, including coloc^25^, fastEnloc^26^, eCAVIAR^27^, FUSION^28^, summary-based Mendelian randomization (SMR)^29^, and PrediXcan^30,31^ converged on a focused set of genes with consistent support. Restricting our analyses to brain tissues, we prioritized 14 high-confidence genes that met the threshold of at least three supporting approaches. These include *GLO1*, *SKOR1, MAP2K5, HOXB*2, and *HOXB3,* which have been repeatedly reported in earlier GWASs, as well as additional candidates like *OMA1*, *ACP1, ALKAL2, TSPOAP1, LRRC45* that showed strong support across multiple brain tissues (Table S15).

### Evaluation of gene druggability

DrugBank searches indicated that several prioritized genes have reported associations with compounds at different stages of development. Approved compounds were identified for multiple genes, including *TACSTD2*, *ACVR1, RARB, TACR3, HEXB, PAM, GLO1, MGMT, BDNF, GRIA4* and *CDH11*. These associations reflect known gene-compound relationships rather than condition-specific targeting (Table 4).

**Table 4.** Druggability of genes identified by MAGMA gene-based analysis based on DrugBank annotations. Approved drugs: drugs that have been officially accepted for commercialization in at least one jurisdiction at a given time; Investigational drug: the drug development status in which the drug is being researched for a determinate condition and has reached clinical trials; Nutraceutical drugs: drugs which are regulated and processed at a pharmaceutical grade and have a demonstrable nutritional effect; Experimental drug: also referred as a drug in discovery or pre-discovery phase, alludes to the drug development status in which the drug is being researched pre-clinically.

### Genetic correlation across European cohorts

To evaluate consistency of phenotype definition across datasets, we estimated pairwise genetic correlations (r_g_) between the European-ancestry cohorts (Table S4). Overall, substantial genetic overlap was observed across studies. The highest concordance was detected between the EHR-based cohorts All of Us and MVP (r_g_ = 0.86, SE 0.07), and the UK Biobank questionnaire-defined cohort also showed strong similarity with these datasets ((r_g_ = 0.78-0.82). The CLSA cohort demonstrated lower correlations with other cohorts (r_g_ = 0.64-0.79). When compared with previously published European GWAS summary statistics (EU-RLS-GWAS), the MVP cohort showed the strongest agreement (r_g_ = 0.92).

### Polygenic prediction in an independent target cohort

Polygenic risk scores (PRS) were evaluated in an independent European ancestry cohort from All of Us (441 cases and 12,172 controls) that was not included in the discovery GWAS. Four independent training GWAS summary statistics were evaluated: (i) the European multi-cohort meta-analysis from the current study (All of Us WGS, MVP, UK Biobank, CLSA, CARTaGENE); (ii) the clinically ascertained EU-RLS-GENE GWAS reported by Schormair et al., 2024^13^; (iii) a survey-based UK Biobank GWAS; and (iv) an electronic health record-based All of Us GWAS from the current study.

Across all training datasets, higher PRS was significantly associated with RLS status after adjustment for age, sex, and PCs (Table S16, Table S17). The PRS derived from the European multi-cohort meta-analysis (PRS_current_EUR) showed the strongest performance, with an odds ratio of 1.78 per SD increase (95% CI 1.62-1.97, P=5.1×10^-31^), improving discrimination from AUC 0.57 to 0.68 (ΔAUC=0.10) when evaluated in an independent All of Us target cohort of European ancestry participants not included in the GWAS meta-analysis (Figure S7). In comparison, PRS trained on the clinically ascertained EU-RLS-GENE cohort (PRS_EU-RLS-GENE) yielded a smaller effect (OR=1.34, 95% CI 1.22-1.48; ΔAUC=0.035), while PRS based on UK Biobank survey phenotypes showed an OR of 1.41 (ΔAUC=0.048), whereas the PRS derived from All of Us EHR phenotypes showed an OR of 1.40 (ΔAUC=0.040) (Table S16). Given the substantial case-control imbalance in the target cohort (441 cases and 12,172 controls), we additionally evaluated model performance using precision-recall curves (Figure S7c), which yielded conclusions consistent with ROC-based evaluation.

To evaluate patient-level risk enrichment, we compared individuals in the highest versus lowest PRS decile. For PRS_current_EUR, individuals in the top decile had six-fold higher odds of RLS compared with the bottom decile (OR=6.01, 95% CI 3.63-9.94, P=2.9 × 10^-12^). The corresponding contrasts were more modest for PRS_EU-RLS-GENE (OR=3.19), PRS_survey-based_UKB (OR = 2.47), and PRS_EHR-based_AoU (OR=3.63) (Table S16). These findings demonstrate a clear genetic risk gradient, with the meta-analysis-derived PRS providing the greatest separation between high and low-risk groups.

## Methods

### Participants and study design

The current genome-wide association meta-analysis included two African ancestry, two Latin American ancestry, and five European ancestry cohorts (Table S1). For the African and Latin American ancestry groups, we utilized data from the All of Us Research Program^32^ and the Million Veteran Program^19^. For the European ancestry group, we incorporated data from All of Us, UK Biobank^33^, the Canadian Longitudinal Study on Aging (CLSA)^34^, CARTaGENE^35^, and the Million Veteran Program^19^. GWAS in the All of Us and UK Biobank cohorts were conducted in the present study using individual-level data, whereas previously published summary statistics were used for the remaining cohorts.

RLS phenotype was defined using EHR and RLS diagnostic questionnaires (Table S2). All contributing studies were approved by relevant Institutional Review Boards and all participants provided informed consent. Detailed descriptions of informed consent procedures, ethical approvals, data governance frameworks, and study protocols for each cohort are provided in the Supplementary Material. A detailed breakdown of the included cohorts and participant numbers is provided in Figure 1 and Table S1.

Across cohorts, participants were drawn from large population-based and healthcare-linked studies conducted primarily in the United States, the United Kingdom, and Canada. Accordingly, the included samples largely represent individuals who are engaged with healthcare systems rather than a random sample of the general population. Heterogeneity in case ascertainment reflects differences in study design and healthcare context across cohorts and may influence generalizability, particularly for individuals with undiagnosed or untreated RLS.

### Sequencing and genotyping procedures

Genotyping and quality control procedures for the CLSA, CARTaGENE, and Million Veteran Program datasets were performed as described in previously published studies^9,19,36^. Additional details on these external datasets are provided in the Supplementary Methods.

Whole-genome sequencing (WGS) for the All of Us Research Program was performed on Illumina NovaSeq 6000 sequencing instruments using PCR-free library preparation, and data were processed through the DRAGEN pipeline (Illumina, San Diego, CA, USA) with multiple layers of quality control before joint variant calling across all samples. Sequence reads were aligned to the GRCh38DH reference genome, and standardized procedures minimized batch effects and yielded uniform sequencing data with an average coverage >= ×30. All of Us samples were generated using WGS and were filtered according to the standard All of Us sequencing quality control pipeline. Samples were required to have a mean coverage of 30× and to pass the All of Us sample-based quality control. Samples were excluded if they showed evidence of erroneous sequencing signals, defined as fewer than 2.4 million or greater than 5.0 million SNPs, more than 100,000 variants absent from gnomAD v3.1, or a heterozygous to homozygous ratio greater than 3.3 for either SNPs or indels. Only samples passing these criteria were included in downstream analyses^32^. Genetic ancestry estimation, relatedness assessment, and principal component analysis were conducted using Hail within the All of Us Researcher Workbench. Ancestry assignment was performed using projection-based methods onto reference datasets including the 1000 Genomes Project and the Human Genome Diversity Project (HGDP). Relatedness was evaluated based on kinship coefficients and KINSHIP[>[0.0884, corresponding to the lower boundary of second-degree relatedness, were removed through pairwise filtering, retaining one individual per related pair.^32^.

Sequencing on individuals for UK Biobank was carried out on the Illumina NovaSeq 6000 platform, generating data with a mean depth of coverage of 34x. Reads were aligned to the GRCh38 reference genome and analyzed using DRAGEN v.3.7.8 (Illumina, San Diego, CA, USA). Further details on quality control can be found at https://biobank.ndph.ox.ac.uk/showcase/label.cgi?id=187^33^. Genetic ancestry was determined using a custom ancestry prediction pipeline implemented in the GenoTools package^37^. Samples with a call rate below 95% were removed. Relatedness was calculated with KING (v2.2.8), following the guidelines provided in the KING manual (available at https://www.kingrelatedness.com/manual.shtml), and individuals closer than cousins (KINSHIP[>[0.088) were pruned using pairwise filtering, retaining one individual from each related pair.

### Genome-wide association analyses

Genome-wide association analyses of RLS case-control status were performed using standard GWAS methodology and quality-control procedures. Under the additive genetic model, genotypes were coded as 0, 1, or 2 according to the number of effect alleles, with the model estimating the change in risk per additional allele. Age at onset and symptom frequency were examined in case-only analyses among RLS cases for variants reaching genome-wide significance. Downstream follow-up analyses were considered exploratory and hypothesis-generating. GWAS were conducted for the All of Us and UK Biobank cohorts in the present study using participant-level case-control cohorts. Association testing was carried out using PLINK v2^23^, adjusting for age, sex, and the first ten principal components (PCs). For cohorts analyzed using previously published summary statistics, covariate adjustment followed the analytical frameworks defined in the original studies. In the MVP GWAS, ten PCs were included based on evaluation of scree plots to capture population structure^19^. For the CLSA and CARTaGENE cohorts, the number of PCs were selected using a stepwise procedure. PC2 and PC6 were included in the CLSA study and PCs 1, 2, 4, 7, 10 were included in the CARTaGENE study^9^. For the All of Us analyses, PCs were calculated within each ancestry group and for the combined cohort (Figure S1a). Scree plots show that the first PCs capture major structure, with PCs 4-10 providing additional, smaller contributions that help adjust for remaining variation in the data (Figure S1b). For UK Biobank, we used the PCs provided by the Genotools pipeline^37^, and ten PCs were included following the approach used in the All of Us analyses and the MVP GWAS^9,19^. Logistic regression was used to analyse the binary case-control outcome, and independence of observations was ensured by excluding related individuals (second degree or closer relatives) prior to analysis. Genome-wide significance was defined as P < 5 × 10^-8^ to account for multiple testing across the genome. Quality-controlled variants with a minor allele frequency >= 0.01, variant missingness < 5%, and Hardy-Weinberg equilibrium P > 1 × 10^-6^ in controls were included in the downstream analysis.

Variant-based heritability (h^2^) and genetic correlation was estimated using LD Score Regression (LDSC) (v1.0.1)^38^. Summary statistics for each cohort and ancestry-specific meta-analysis were harmonized using munge_sumstats.py, excluding variants with a minor allele frequency below 0.01. Ancestry-matched LD reference panels derived from the UK Biobank were used for the European, African, and Latin American analyses^39^. LDSC was applied for each cohort and meta-analysis to estimate liability-scale heritability, adjusting for case-control ratios.

### Ancestry-stratified GWAS meta-analyses

To provide context for the ancestry-specific genome-wide association analyses, we evaluated statistical power across a range of allele frequencies and genotype relative risks using cohort-specific sample sizes under an additive genetic model in each ancestry.

We estimated statistical power using the Genetic Association Study (GAS) Power Calculator (https://csg.sph.umich.edu/abecasis/cats/gas_power_calculator/). Power calculations were performed under an additive genetic model assuming a one-stage case-control GWAS design and a genome-wide significance threshold of 5 × 10^-8^[. For African and Latin American ancestry analyses, we used the observed proportion of RLS cases in the All of Us cohort (0.009) as a proxy for disease prevalence, as reliable population-based prevalence estimates for these ancestry groups are not currently available. For European ancestry analyses, a prevalence of 0.06 was used, consistent with estimates from the UK Biobank and Canadian cohorts. Power was evaluated across a range of allele frequencies (0.01-0.25) and odds ratios (1.2, 1.5, and 2.0).

Ancestry-stratified meta-analyses were performed using an inverse-variance weighted fixed-effects model implemented in METAL^40^ and a random-effects model implemented in PLINK v2 to account for potential between-study heterogeneity. For the African and Latin American ancestry meta-analyses, overlapping variants between the All of Us and Million Veteran Program datasets were included. For the European meta-analysis, which combined five cohorts, variants present in at least three datasets were analyzed. To account for the imbalance between case and control sample sizes, genomic inflation factors (λ) were normalized to a standard sample size of 1,000 cases and 1,000 controls using the formula λ_1000_ = 1 + (λ − 1) × (1/number of cases + 1/number of controls) × 500^41^. Independence of the reported loci was assessed using LD-based comparisons within the ancestry-specific reference panels using the LDpair tool (https://ldlink.nih.gov/ldpair). Variants with R^2^ < 0.1 with known lead variants and separated by >500 kb were considered independent. Additionally, we performed conditional and joint analysis using GCTA-COJO (v1.94.3) to identify conditionally independent association signals within each locus. For each meta-analysis, LD was estimated from the corresponding ancestry subset of the 1000 Genomes Project Phase 3 reference panel aligned to GRCh38. Independent genome-wide significant signals were merged into a single genomic risk locus if their LD blocks were separated by less than 500 kb.

### Multi-ancestry meta-analysis

Summary statistics from nine GWASs representing two African-ancestry, two Latin American-ancestry, and five European-ancestry cohorts were included. Association summary statistics were aggregated using two complementary approaches: (i) inverse-variance weighted fixed and random-effects models implemented in PLINK v2^23^, and (ii) a multi-ancestry meta-regression implemented in MR-MEGA^22,23^. The PLINK analyses were used to obtain conventional pooled effect estimates and measures of between-study heterogeneity (I^2^), consistent with standard practice in GWAS meta-analyses^42,43^. MR-MEGA was applied to combine cohorts while accounting for ancestry differences through ancestry principal components derived from study-level allele frequency variation, with the aim of improving detection of shared association signals across populations. Variants present in at least three datasets were retained for downstream analyses. Genome-wide significance was defined as P < 5 × 10^-8^ to account for multiple testing across the genome.

### *APOE*-specific analyses

We assigned *APOE* diplotypes from rs429358 and rs7412 and annotated individuals into ε2/ε3/ε4 genotype groups, excluding those with missing or ambiguous calls. We tested associations with RLS using three logistic regression models: an ε2-additive model, an ε4-additive model, and a genotype model with ε3/ε3 as the reference. We adjusted all models for age, sex, and the first ten principal components. These analyses were performed only in cohorts with individual-level genotype data (UK Biobank and All of Us ancestry-specific datasets).

To evaluate the robustness of the *APOE* association, we additionally performed an age-restricted sensitivity analysis in the UK Biobank by repeating the case-control GWAS after restricting the analysis to participants younger than 60 years at recruitment (cases = 2,446, controls = 35,896). The same regression framework and covariates were used as in the primary analysis.

### Association of lead variants with frequency of symptoms and age at onset

We further evaluated associations between lead variants and age at onset and frequency of symptoms using individual level data from the All of Us and UK Biobank, respectively. We evaluated associations between lead variants and symptom frequency in the UK Biobank, where information on RLS symptom frequency was available. We defined RLS cases based on diagnostic questionnaire items (Table S2) and restricted analyses to individuals reporting symptoms occurring at least 2-3 days per week. Symptom frequency was assessed using self-reported responses describing how often RLS symptoms occurred over the previous 12 months. We included only cases with available symptom frequency data (n = 3,380), comprising individuals reporting symptoms on 2-3 days per week (n = 1,597), 4-5 days per week (n = 989), or 6-7 days per week (n = 794). We performed case-only analyses and modeled symptom frequency as an ordinal outcome with three levels using a proportional-odds framework, adjusting for age, sex, and the first ten PCs. Statistical significance was assessed using a Bonferroni-corrected threshold accounting for 81 lead variants identified in the European meta-analysis (P < 0.0006, α = 0.05 / 81).

We evaluated associations between lead variants and age at onset using individual-level data from the All of Us cohort across African, Latin American, and European ancestry groups, as age at onset was not available in the UK Biobank cohort. Analyses were restricted to RLS cases with available age at onset data and were tested using linear regression models adjusted for sex and the first ten PCs. A Bonferroni correction was applied to account for multiple testing across the lead variants in 83 loci identified in the ancestry stratified meta-analysis.

### Fine-mapping analysis

We performed cross-ancestry fine-mapping using SuSiEx, which integrated summary statistics and ancestry-specific LD information to identify potential causal variants^24^. SuSiEx enables joint fine-mapping across populations with distinct linkage disequilibrium (LD) patterns, thereby improving resolution and prioritization of variants underlying association signals. SuSiEx derives posterior inclusion probability by evaluating how well each variant explains the association signal relative to neighbouring correlated variants, so variants that provide information beyond what is captured by LD receive higher probabilities.The PIP is therefore a value between 0 and 1 that reflects how likely a variant is to be causal for the observed signal at a given locus. The analysis included African, Latin American, and European ancestry meta-analyses. For each genome-wide significant locus, we defined a 1 Mb region centered on the lead variant and generated ancestry-specific LD reference files from All of Us individual-level data using PLINK v2. SuSiEx was executed jointly across ancestries, incorporating summary statistics, sample sizes, and LD matrices for each population. Fine-mapping was performed using default parameters, with a 0.95 credible set threshold, and a p value threshold of 1[×[10^-5^. Variants were filtered at PIP > 0.8, in line with the threshold used in the original SuSiEx study to denote high-confidence candidates^24^.

### Cross ancestry gene-based tissue-specific enrichment and colocalization analyses

Gene-based and tissue enrichment analyses were conducted using MAGMA v1.08 implemented in FUMA v1.5.2^44^. Summary statistics from ancestry-specific meta-analyses and from the random effect multi-ancestry GWAS were used as input. LD structure was estimated using ancestry-matched reference panels available within FUMA, derived from the UK Biobank (for European GWAS), and 1000 Genomes Project Phase 3 African and American datasets. This approach combines SNP-level GWAS results to evaluate each gene as a whole rather than testing variants one by one. MAGMA considers all SNPs within a gene together and accounts for the correlations between them caused by LD, so that correlated variants do not artificially inflate the signal. The method produces a single gene-level test statistic and p-value that summarize the overall evidence for association of that gene. The analysis was based on GWAS summary statistics rather than individual-level data. MAGMA gene-based tests were applied to all annotated genes to evaluate aggregated variant-level associations. Statistical significance for gene-based association tests was determined using Bonferroni correction based on the number of genes tested within each ancestry-specific analysis (European analysis: p < 2.63 × 10^-6^, α = 0.05 / 18,987; Latin American analysis: P < 2.60 × 10^-6^, α = 0.05 / 19,232; multi-ancestry random-effect meta-analysis: p < 4.80 × 10^-6^, α = 0.05 / 10,412).

Tissues-specific expression enrichment was examined using GTEx v8 transcriptomic data across 53 tissues. All analyses were conducted using default FUMA parameters and Ensembl v102 gene annotations^44^. This method examines the association between gene-level GWAS signals and gene expression levels from GTEx to determine whether genes that are highly expressed in a specific tissue show stronger genetic association than other genes. The analysis adjusts for gene size, SNP density (the number of SNPs mapped to each gene), and LD to reduce bias.

We used LocusCompare2 (https://locuscompare2.com/) to evaluate locus-level correspondence between association signals across ancestries by integrating GWAS results with gene-expression-based datasets^31^. Ancestry-stratified summary statistics from the African, Latin American, and European meta-analyses were uploaded to the platform. Linkage disequilibrium was calculated using ancestry-matched 1000 Genomes Project Phase 3 reference panels (AFR, AMR and EUR, respectively), as implemented within the tool. Six colocalization and transcriptome-integration tools, including coloc^25^, fastEnloc^26^, eCAVIAR^27^, FUSION^28^, summary-based Mendelian randomization (SMR)^29^, and PrediXcan^30^, were run through LocusCompare2 using GTEx v8-derived expression weights across 14 brain tissues and whole blood. Within LocusCompare2, SMR analyses were carried out using SMR v1.3.0^29^, which performs both the SMR and HEIDI tests. GTEx v8 eQTL summary statistics were used as the molecular QTL reference. Genes were required to have at least one cis-eQTL at P < 1×10^-5^. For the HEIDI test, LD was estimated from ancestry-matched 1000 Genomes Project Phase 3 reference panels in PLINK format, as implemented in the platform. Alleles were harmonized across GWAS, eQTL and LD reference datasets prior to analysis. To ensure comparable stringency across methods, we applied the following thresholds: coloc posterior probability H4 >= 0.75, fastENLOC GRCP >= 0.5, and eCAVIAR CLPP >= 0.01. For transcriptome-based approaches (PrediXcan, FUSION and SMR), we used a p value cutoff of 2×10^-6^, reflecting a Bonferroni correction for approximately 20,000 genes tested per GTEx brain tissue.

### Druggability of prioritized genes

To assess potential druggability, we queried the DrugBank database (https://go.drugbank.com/) for reported gene-compound associations^45^. Genes identified by the MAGMA gene-based analysis were examined and associated compounds were grouped into four categories according to DrugBank annotation: approved, investigational, nutraceutical, and experimental. No filtering based on indication or therapeutic relevance was applied.

## Discussion

We present the first large-scale, multi-ancestry GWAS of RLS, incorporating two African ancestry, two Latin American ancestry, and five European ancestry cohorts. By combining individual-level analyses with harmonized summary statistics, we broadened the genetic landscape of RLS beyond European ancestry and uncovered novel ancestry-specific and shared risk loci. We identified two novel loci near *PRIMA1* and *GYPC/TEX51* in the African ancestry GWAS and one near *ISX* in the Latin American GWAS, in addition to 11 novel loci in the European ancestry analysis. In the combined multi-ancestry meta-analysis, we detected ten novel RLS risk loci, seven of which overlapped with signals from the European GWAS, while three loci reached genome-wide significance exclusively in the multi-ancestry analysis.

Among the identified loci, *SKOR2* is of particular interest, as it is a paralog of *SKOR1*, one of the most consistently replicated genetic signals in RLS^9,13,14^. While earlier GWAS have repeatedly highlighted the neighboring *SKOR1-MAP2K5* region on chromosome 15, our identification of *SKOR2* on a different chromosome suggests that more than one member of the *SKOR* family may contribute to RLS risk. *SKOR1* encodes a SKI-family transcriptional corepressor expressed in central nervous system neurons and involved in neuronal differentiation programs^46,47^. Prior functional work demonstrated that *MEIS1*, a major RLS risk gene, regulates *SKOR1* expression through binding at the *SKOR1* promoter, and that RLS-associated variants modify this regulatory interaction, thereby prioritizing *SKOR1* over the neighboring *MAP2K5* gene within the associated locus^48^. Experimental studies indicate that *SKOR2*, a transcriptional regulator, is involved in cerebellar Purkinje cell differentiation and maturation, consistent with a role in neuronal developmental pathways^49^. In this context, our identification of *SKOR2*, a paralog of *SKOR1*, suggests that multiple members of the *SKOR* family may be involved in transcriptional programs relevant to RLS risk. Consistent with this theme, *MEIS2*, which encodes the homeobox transcription factor Meis2 and is a paralog of *MEIS1*, was previously implicated in functional enrichment analyses of RLS GWAS loci^14^. The convergence of genetic association at *SKOR2* with prior *MEIS1-SKOR1* regulatory evidence supports further investigation of shared regulatory or developmental pathways in RLS.

*RARB* represents another transcriptional regulator identified in our analyses*. RARB* encodes retinoic acid receptor β, a ligand-dependent transcription factor mediating retinoid signaling during neural development. Previous studies showed that loss of retinoic acid receptor β function disrupts striatonigral projection neuron differentiation within the lateral ganglionic eminence, is accompanied by altered Meis1 expression in the developing basal ganglia, and is associated with reduced expression of dopamine receptor D in striatonigral neurons^50,51^. These findings place *RARB* and *MEIS1* within related developmental programs regulating striatal neuronal identity and aspects of dopaminergic signaling.

The *MEIS1* and *BTBD9* loci, two of the most consistently replicated RLS risk loci in European cohorts, were also detected in our Latin American meta-analysis but did not reach significance in the African ancestry analysis, despite similar sample size between two datasets. This difference is largely driven by allele frequency rather than a true absence of effect. The lead *MEIS1* variant rs113851554-T is approximately five times less common in African ancestry populations than in Europeans (gnomAD v4.1), and the lead *BTBD9* variants rs9369062-C in the Latin American analysis and rs4714163-C in the European analysis show a similar reduction in frequency in African ancestry. The reduced frequencies of these variants may explain some of the reduced prevalence in African-ancestry populations^52,16,53^. Notably, studies conducted in East Asian cohorts have also not replicated the *MEIS1* association^54,55^. However, observed differences in RLS prevalence across ancestry groups may also reflect differences in diagnosis, healthcare access, and phenotypic ascertainment, and should not be attributed solely to genetic risk allele frequencies. At the same time, the emergence of distinct genome-wide significant signals in the African ancestry GWAS suggests that genetics of RLS differ across populations, and that risk in non-European groups may be shaped by additional or ancestry-specific variants. Power analyses highlight limited sensitivity to detect modest effect sizes in African and Latin American ancestry analyses compared to European ancestry. This difference reflects sample size disparities across ancestry groups and should be considered when interpreting ancestry-specific results.

The *MEIS1* rs113851554-T allele, a well-established common susceptibility factor for RLS, was associated with a later age at onset in a dosage-dependent manner. This finding highlights that genetic susceptibility and disease timing represent distinct aspects of RLS, as variants that increase lifetime risk do not necessarily lead to earlier symptom onset. Rather than implying a causal delay of disease onset, this association is best interpreted as a higher representation of the *MEIS1* risk allele among individuals with later-onset RLS. In this context, it is also noteworthy that the cohorts analyzed are enriched for elderly subjects (see Table S1), which may influence the distribution of age at onset.

An inverse association at the *APOE* variant was observed in the European analysis and was largely driven by the UK Biobank cohort, in which RLS was defined using an optional questionnaire. To evaluate whether this reflected a true association or a study-design effect, we conducted a GWAS of participation in the RLS diagnostic survey and identified a strong association at rs429358, whereas other reported RLS loci showed no comparable signal. This pattern is consistent with prior studies showing that participation in optional UK Biobank components, such as the mental health questionnaire^21^, has a heritable component and can influence downstream genetic association results. Variants related to cognitive status may therefore affect completion of diagnostic questions, leading to differential misclassification of cases and controls. In addition, survival or ascertainment bias may also play a role, as *APOE* ε4 is associated with dementia risk and mortality, which could influence who participates in studies and is represented in population-based cohorts. We therefore interpret the *APOE* finding cautiously and do not consider it evidence of a protective effect on RLS risk, but rather an effect related to participation in questionnaire-derived phenotypes.

MR-MEGA allowed us to evaluate whether RLS risk loci showed consistent effect sizes across cohorts or evidence of ancestry-dependent variation. In our multi-ancestry meta-analysis, 16 loci showed significant heterogeneity (P_ancestry_het < 0.01), although the magnitude of this signal varied across loci. The strongest differences across ancestry groups were observed at *PDE9A, TRIB2*, and *CRBN* (P < 5 × 10^-4^), where effect sizes differed in magnitude and, in some cases, direction across ancestry groups. In contrast, the *MEIS1* risk variant showed comparable effect sizes across all three ancestries (Table S5), suggesting that differences in significance reflect variation in allele frequency and statistical power rather than true effect heterogeneity.

Tissue enrichment analysis of the European meta-analysis identified enrichment for genes expressed in the cerebellum and cerebellar hemisphere. While this finding is consistent with previous reports implicating cerebellar involvement in RLS^13^, our results provide independent confirmation within the current dataset. The lack of significant enrichment in the African, Latin American, or multi-ancestry meta-analysis likely reflects reduced statistical power in non-European cohorts and the integration of heterogeneous sample sizes across ancestries, which may attenuate tissue-level enrichment signals.

A major strength of the study is the definition of RLS cases, combining electronic health records (EHR)-confirmed diagnoses with validated diagnostic questionnaires. In All of Us and the Million Veteran Program, RLS diagnoses were obtained directly from structured EHRs, whereas in the UK Biobank and the CARTaGENE, case status was based on standardized questionnaire items aligned with the International Restless Legs Syndrome Study Group (IRLSSG) diagnostic criteria. This helped us define RLS cases more reliably than studies that rely solely on single-item self-report. Earlier European meta-analyses combined clinically confirmed datasets with cohorts such as 23andMe, in which RLS status was inferred from a single self-reported question without diagnostic confirmation. Some of the associations observed in that study, including the *HLA* locus, were not replicated in the clinically ascertained European cohort or in our current analysis, suggesting that the signal may have resulted from imprecise phenotyping, as well as limited statistical power in individual cohorts to detect modest effects. Overall, the use of clinically defined or diagnostically validated cases reduces phenotyping noise and improves the interpretability and reproducibility of genetic findings. Differences between studies, however, cannot be attributed to diagnostic approach alone. Heritability estimates varied across cohorts, with the highest estimate observed in the UK Biobank despite its questionnaire-based case definition, whereas genetic correlation analyses showed closer agreement between the EHR-based cohorts and the clinically ascertained EU-RLS-GWAS dataset. Nevertheless, these analyses provide a quantitative link between genetic findings and observable disease variation and could, with further replication and validation, contribute to future clinical trial stratification strategies. We included PRS analyses to complement the locus-based GWAS findings with a quantitative assessment of genetic risk. The PRS derived from the multi-cohort meta-analysis, which combined EHR-based and survey-based diagnoses, showed the strongest performance across our evaluations. In comparison, the PRS trained on the clinically ascertained EU-RLS-GENE dataset showed more modest effects, underscoring that PRS performance depends on factors such as sample size, cohort composition, and the alignment between training and target phenotypes. Given that our target cohort was EHR-based and the meta-analysis included substantial biobank-derived data, greater concordance between these settings may have contributed to the observed differences. These analyses provide an empirical link between genetic findings and patient-level variation that can be examined further in future studies.

Although this study represents a step forward in expanding genetic research on RLS beyond European ancestry, several limitations should be acknowledged. First, although this study integrates large-scale genomic data from European ancestry cohorts, sample sizes for the African and Latin American ancestry analyses remain more limited, reflecting the current under-representation of these populations in genomic research. The absence of other large, independent RLS GWAS datasets with substantial African or Latin American representation precluded formal replication of ancestry-specific signals identified in this study. In the MVP dataset, replication was limited to the established *MEIS1* and *BTBD9* loci^19^. Additional associations were identified through integration with previously unpublished All of Us African and Latin American data, highlighting the importance of expanding diverse genomic resources. Across cohorts, participants are drawn primarily from population-based and healthcare-linked biobanks in high-income countries and therefore are not fully representative of the global population. Participants tend to be older and more likely to be female and are typically engaged with healthcare systems. This may under-represent individuals with undiagnosed, untreated, or milder forms of RLS. Heterogeneity in case ascertainment across cohorts reflects differences in study design, healthcare access, and diagnostic practices, and may influence the generalizability of findings across populations. In addition, statistical power to detect modest-effect or low-frequency variants may be reduced in the African and Latin American ancestry analyses due to smaller sample sizes. For example, the *MEIS1* rs113851554 risk allele is less frequent in African ancestry populations, resulting in reduced power to detect association in this locus. The absence of genome-wide significance at this locus in African ancestry GWAS reflect limited power rather than absence of true effect. Larger and more diverse cohorts will be essential to refine effect-size estimates, improve fine-mapping resolution, and ensure the broad applicability of genetic findings in RLS. Second, detailed information on disease stage, symptom severity, and comorbid conditions was not uniformly available across cohorts. Because RLS often co-occurs with other neurological and systemic conditions, and because several cohorts relied on electronic health records or survey-based definitions, some degree of clinical heterogeneity cannot be excluded. This includes the possibility that not all cases represent strictly idiopathic RLS. These factors may influence the generalizability of our findings and should be considered when interpreting effect size estimates across cohorts. Third, we could not perform sex-stratified analyses. The Million Veteran Program only provided sex-combined full summary statistics, and the sample sizes in All of Us and the non-European cohorts were too small to support reliable stratification. As a result, we could not assess whether any of the identified associations differ by sex, despite the well-established sex imbalance in RLS prevalence. Fourth, even though this is the largest genomic study to date in African and Latin American ancestry populations, the sample sizes in these groups are still much smaller than in Europeans, and publicly available RLS GWAS datasets from non-European ancestry populations are limited. Fifth, cross-ancestry fine-mapping was performed using LD reference panels constructed from individual-level All of Us genome data, which offered both sufficient scale and diverse ancestry representation. However, because the meta-analysis includes cohorts with different admixture profiles, LD patterns in All of Us may not fully capture the haplotype structure of every dataset included in the analysis. In addition, LocusCompare2 and related colocalization analyses relied on GTEx gene expression data, which are predominantly derived from individuals of European ancestry. Although LD structure was ancestry-specific, the expression reference was not, and ancestry-specific regulatory effects may therefore be incompletely captured. Finally, variant-based heritability could not be reliably estimated in the African and Latin American ancestry cohorts due to limited case numbers and the lack of comprehensive ancestry-specific LD reference panels, leading to unstable or non-convergent LDSC models. Nevertheless, the genomic inflation factors in these analyses were close to 1, indicating well-calibrated association statistics.

In conclusion, this first large-scale genomic study of RLS across multiple ancestries identifies two novel loci in African ancestry, one in Latin American ancestry, eleven in European, and ten additional loci through multi-ancestry meta-analysis, including three loci uniquely identified in the multi-ancestry analysis and seven that overlapped with novel European signals. By analyzing clinically defined RLS cases across diverse populations, we refine the genetic understanding of the disorder and demonstrate that broader ancestral representation both enables the discovery of ancestry-specific loci and reveals which associations are shared across populations. Continued expansion of diverse cohorts will be essential for ensuring that future genetic findings are transferable and informative for RLS biology and, ultimately, for clinical management.

## Data and code sharing

Publicly available data consortia include All of Us Research Program (https://www.researchallofus.org/register/) and UK Biobank (https://www.ukbiobank.ac.uk/enable-your-research/register), in which data can be accessed after applying. A repository of the code used for processing and analyzing is publicly available (https://github.com/fulyaakcimen/fulyaakcimen-Multi-ancestry-GWAS-for-RLS/). CLSA data are available from the Canadian Longitudinal Study on Aging (www.clsa-elcv.ca) for researchers who meet the criteria for access to de-identified CLSA data.

## Data Availability

https://github.com/fulyaakcimen/fulyaakcimen-Multi-ancestry-GWAS-for-RLS/

## Acknowledgments

We thank Suleyman Can Akerman and Paige Jarreau for their meticulous editing of this manuscript and figures. This research was supported, in part, by the Intramural Research Program of the National Institutes of Health (NIH). The contributions of the NIH author(s) are considered Works of the United States Government. The findings and conclusions presented in this paper are those of the author(s) and do not necessarily reflect the views of the NIH or the U.S. Department of Health and Human Services. This work utilized the computational resources of the NIH HPC Biowulf cluster. (http://hpc.nih.gov).

The All of Us Research Program is supported by the National Institutes of Health, Office of the Director: Regional Medical Centers: 1 OT2 OD026549; 1 OT2 OD026554; 1 OT2 OD026557; 1 OT2 OD026556; 1 OT2 OD026550; 1 OT2 OD 026552; 1 OT2 OD026553; 1 OT2 OD026548; 1 OT2 OD026551; 1 OT2 OD026555; IAA #: AOD 16037; Federally Qualified Health Centers: HHSN 263201600085U; Data and Research Center: 5 U2C OD023196; Biobank: 1 U24 OD023121; The Participant Center: U24 OD023176; Participant Technology Systems Center: 1 U24 OD023163; Communications and Engagement: 3 OT2 OD023205; 3 OT2 OD023206; and Community Partners: 1 OT2 OD025277; 3 OT2 OD025315; 1 OT2 OD025337; 1 OT2 OD025276. In addition, the All of Us Research Program would not be possible without the partnership of its participants.

This research was made possible using the data/biospecimens collected by the Canadian Longitudinal Study on Aging (CLSA). Funding for the Canadian Longitudinal Study on Aging (CLSA) is provided by the Government of Canada through the Canadian Institutes of Health Research (CIHR) under grant reference: LSA 94473 and the Canada Foundation for Innovation, as well as the following provinces, Newfoundland, Nova Scotia, Quebec, Ontario, Manitoba, Alberta, and British Columbia. This research has been conducted using the CLSA dataset (Identification of genetic loci associated with restless legs syndrome in the Canadian population, CLSA Comprehensive Follow-up 1 dataset version 3.1, and Comprehensive Baseline dataset version 6.1 and Genome-wide Genetic Data – Version 3.0, under Application Number 2104033). The CLSA is led by Drs. Parminder Raina, Christina Wolfson and Susan Kirkland. The time and commitment of the participants to the CLSA study platform is gratefully acknowledged, without whom this research would not be possible. The opinions expressed in this manuscript are the author’s own and do not reflect the views of the Canadian Longitudinal Study on Aging.

This research has been conducted using the UK Biobank Resource under application number 33601.

## Contributors

FA, KLCJ, and EM contributed to the study concept or design. FA, KLJC, GAR, PAD, GAR, and EM were involved in sample and data acquisition and access to raw data. FA, MK, and MM did the analysis. All authors contributed to critical review and had final responsibility for the decision to submit for publication.

---

Table S1. Restless legs syndrome cohort descriptions.

Table S2. RLS phenotyping and case definition across participating cohorts. Cases were identified using either electronic health records (EHRs) or validated diagnostic questionnaires, depending on cohort. RLS diagnoses in the All of Us and MVP cohorts were defined based on clinical codes from EHRs, while in the UK Biobank, CLSA, and CARTaGENE cohorts, case status was determined using questionnaire responses corresponding to the International Restless Legs Syndrome Study Group diagnostic criteria.

Table S3. Expected statistical power across ancestry groups under an additive genetic model. For the African- and Latin American-ancestry analyses, allele frequency estimates were derived from the All of Us ancestry-specific subsets and used as proxies for power calculations. These estimates may not fully represent allele frequencies across all populations included in the respective meta-analyses.

Table S4. Genomic inflation, variant-based heritability estimates and genetic correlation analyses across GWAS datasets for restless legs syndrome. AoU: All of Us, MVP: Million Veteran Program, CLSA: Canadian Longitudinal Study on Aging, CaG: CARTaGENE. AFR: African, AMR: Latin American, EUR: European ancestry. Population prevalence: estimated disease prevalence in the population, Sample prevalence: proportion of RLS cases, λ: genomic inflation factor; λ1000: inflation scaled to 1000 cases and 1000 controls h²: SNP-based heritability estimate; se: standard error. NA:LDSC liability-scale heritability could not be reliably estimated in this cohort due to low case counts. rg: genetic correlation.

Table S5. Summary statistics of 83 independent genome-wide significant variants in 65 loci identified in ancestry-specific meta-analyses for restless legs syndrome.AFR: African, AMR: Latin American, EUR: European ancestry.

Table S6. Cross-ancestry analysis of variants previously reported in European GWAS meta-analysis for restless legs syndrome. AFR: African, AMR: Latin American, EUR: European ancestry. Power calculations were performed under an additive genetic model assuming a one-stage case–control GWAS design and a genome-wide significance threshold of 5 × 10^-8^.

Table S7. Analysis of *APOE* alleles in restless legs syndrome

Table S8. Genome-wide significant loci associated with participation in the UK Biobank RLS diagnostic questionnaire

Table S9. Association of lead variants with age at onset among RLS cases in the All of Us cohort, stratified by ancestry. AFR: African, AMR: Latin American, EUR: European ancestry.

Table S10. Association of lead variants with symptom frequency among RLS cases in the UK Biobank. Associations between lead variants and symptom frequency were evaluated among UK Biobank cases of European ancestry with available symptom frequency data (n = 3,380; 2-3 days/week, n = 1,597; 4-5 days/week, n = 989; 6-7 days/week, n = 794). Symptom frequency was modeled as an ordinal outcome using proportional-odds regression adjusted for age, sex, and the first ten PCs.

Table S11. Novel genome-wide significant associations in individual ancestry restless legs syndrome cohorts.

Table S12. MR-MEGA multi-ancestry meta-analysis results of lead variants associated with restless legs syndrome. p-value of the heterogeneity due to different ancestry; lnBF:log of Bayes factor.

Table S13. Random effects multi-ancestry meta-analysis results of lead variants associated with restless legs syndrome. Random-effects meta-analysis was performed in PLINK v2. N:number of samples; I2: heterogeneity index (0-100)

Table S14. Gene-based association test results. Genes reaching Bonferroni-corrected significance are listed (European analysis: p < 2.63 × 10-6, α = 0.05 / 18,987; Latin American analysis: p < 2.60 × 10-6, α = 0.05 / 19,232; Multi-ancestry random-effect meta-analysis: p < 4.80 × 10-6, α = 0.05 / 10,412).

Table S15. Putative causal genes. Prioritized genes (supported by at least three evidence counts across the six integrative gene-mapping approaches) are indicated by asterisks (*). Significance thresholds were as follows: coloc posterior probability H4 >= 0.75, fastENLOC GRCP >= 0.5, and eCAVIAR CLPP >= 0.01. For transcriptome-based approaches (PrediXcan, FUSION and SMR), we applied a p-value cutoff of 2×10^-6^, reflecting a Bonferroni correction for approximately 20,000 genes tested per GTEx brain tissue.

Table S16. Performance of polygenic risk scores in the independent target cohort. Odds ratios represent the change in odds of RLS per one standard deviation increase in PRS, estimated from logistic regression adjusted for age, sex, and the first ten PCs. AUC_base indicates discrimination of the covariate-only model; AUC_prs indicates discrimination after addition of PRS; ΔAUC represents the improvement in AUC attributable to PRS. Training sources: PRS_current_EUR:European multi-cohort meta-analysis from the current study; PRS_EU-RLS-GENE: Clinically-ascertained European GWAS (Schormair et al., 2024); PRS_survey-based_UKB: UK Biobank questionnaire-based GWAS; PRS_EHR-based_AoU: All of Us electronic health record-based GWAS.

Table S17. Risk enrichment across PRS deciles in the target cohort. Comparison of individuals in the highest (10th) versus lowest (1st) PRS decile in the independent All of Us target cohort. Odds ratios (OR) were estimated from logistic regression adjusted for age, sex, and the first ten genetic principal components. Training sources: PRS_current_EUR:European multi-cohort meta-analysis from the current study; PRS_EU-RLS-GENE: Clinicaly-ascertained European GWAS (Schormair et al., 2024); PRS_survey-based_UKB: UK Biobank questionnaire-based GWAS; PRS_EHR-based_AoU: All of Us electronic health record-based GWAS.

**Figure S1.**
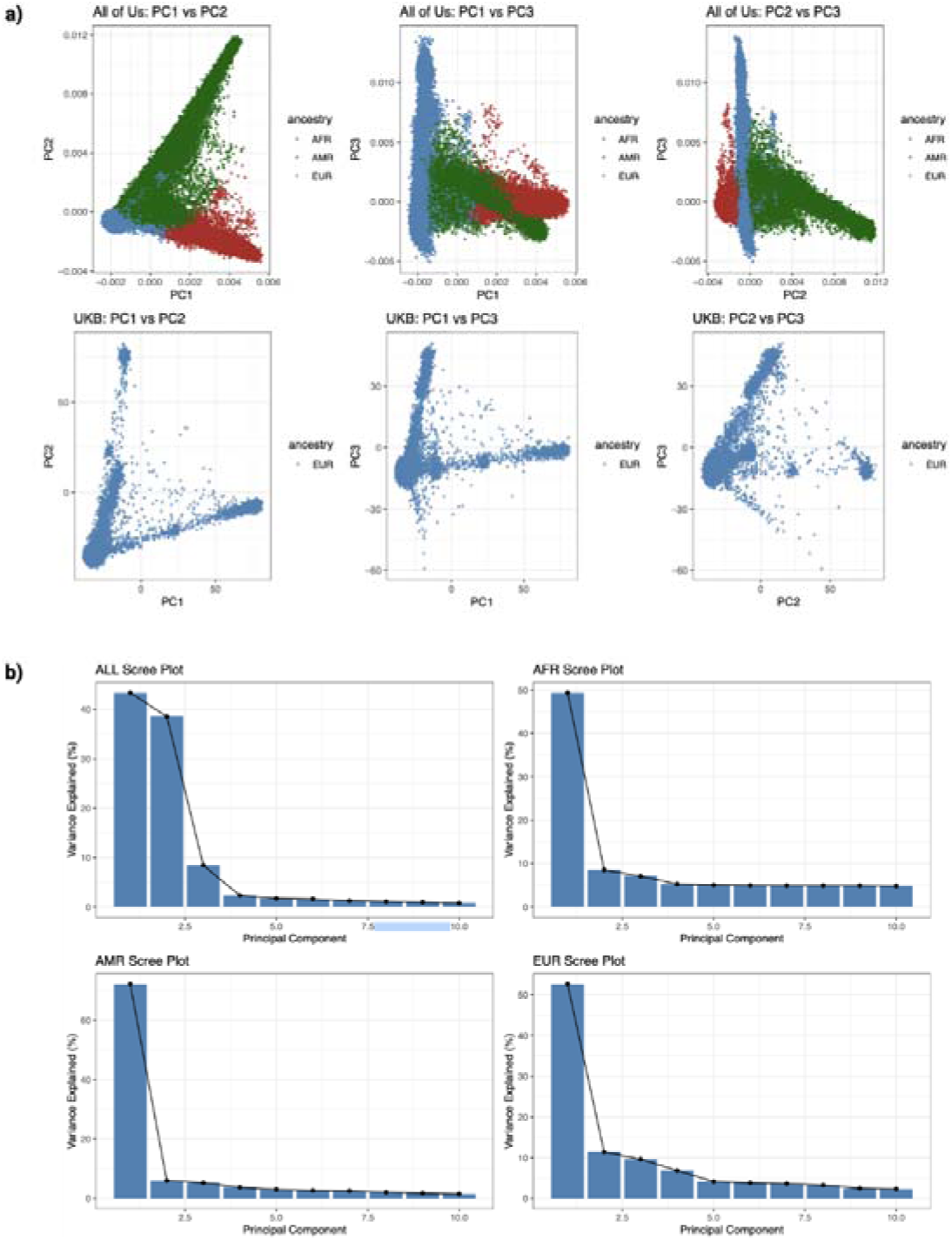
Principal component structure in All of Us and UK Biobank a) Scatter plots of the first three principal components (PC1-PC3) illustrating genetic ancestry structure. b) Scree plots displaying the proportion of variance explained by the first ten principal components for the combined cohort and within each ancestry group in All of Us (AFR, AMR, EUR).

**Figure S2.**
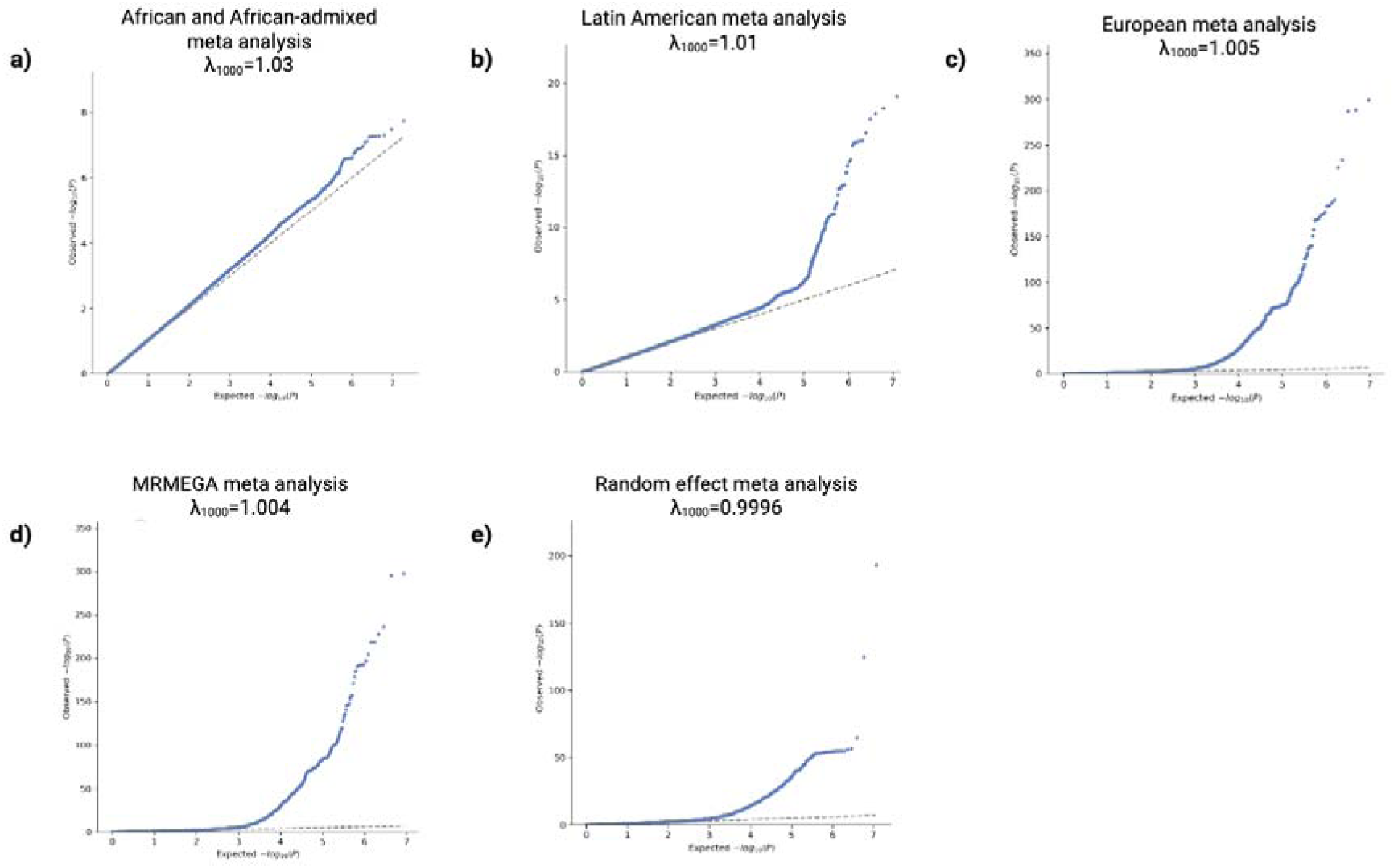
Quantile-quantile (QQ) plots and genomic inflation (λ1000) for each analysis Genomic inflation values were normalized to 1000 cases and 1000 controls for all datasets to account for the large imbalance between case and control sample sizes.

**Figure S3.**
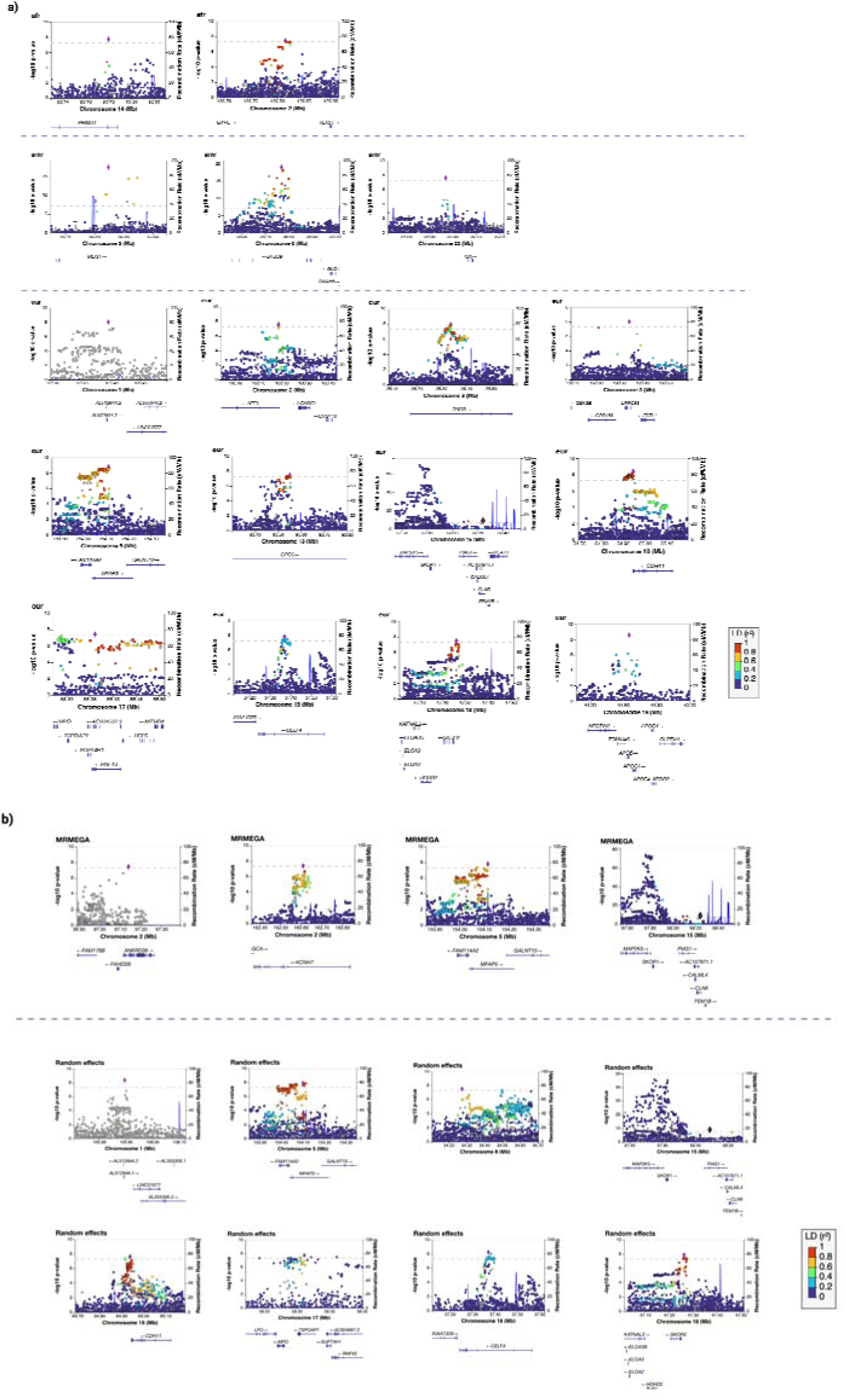
a. LocusZoom plots for new loci identified in ancestry -stratified meta-analyses. b. LocusZoom plots for new loci identified in multi-ancestry MRMEGA or random effect meta-analyses. c. Regional association plots for European (EUR) and Admixed American (AMR) analyses are shown using alternative LD reference variants. d. Regional association plots for the BTBD9 locus shown across EUR, AMR, and AFR ancestry analyses. The LD color scale (bottom right) indicates the strength of linkage disequilibrium (r²) between each variant and the lead SNP, ranging from low LD (blue) to high LD (red).

**Figure S4.**
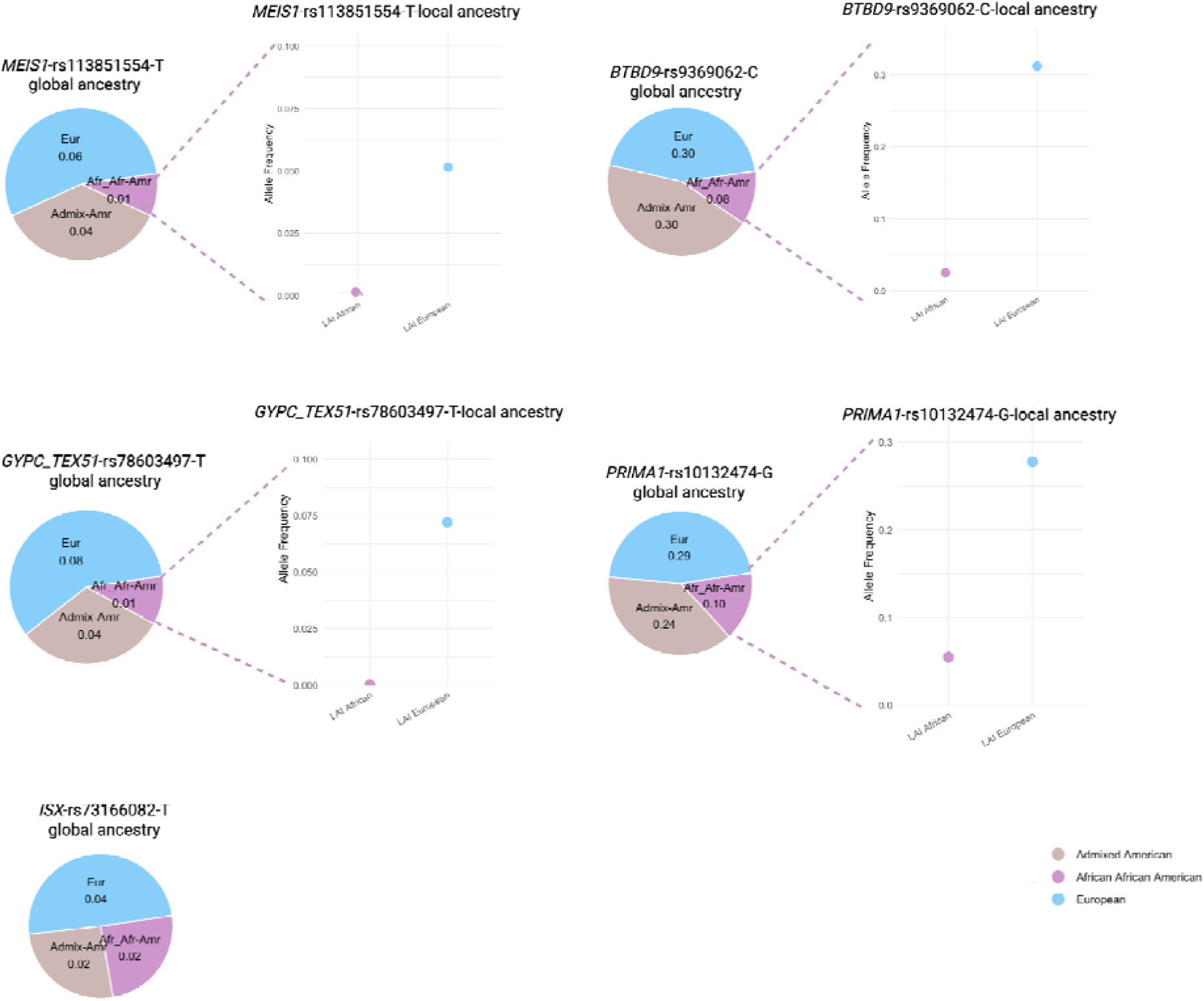
Global and local ancestry allele frequency distribution of the RLS risk variants identified in African and African admixed and Latin American cohorts: *MEIS1, BTBD9, GYPC/TEX51, PRIMA1*, and *ISX*. Local ancestry inference (LAI) for African or African American and Admixed American ancestry groups was obtained from gnomAD v4.1. LAI for the *ISX* lead variant was unavailable.

**Figure S5.**
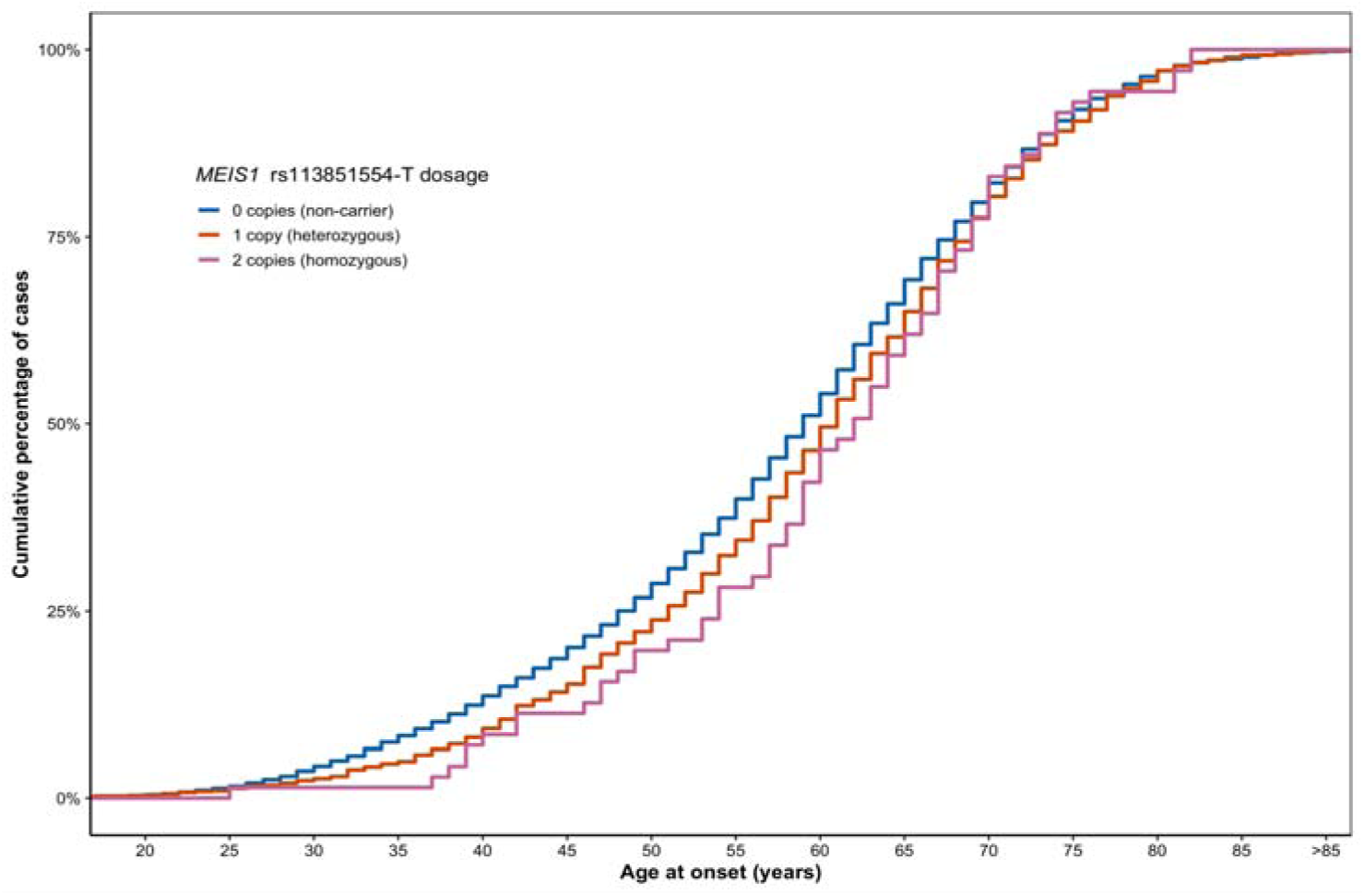
Age at onset of restless legs syndrome according to *MEIS1* rs113851554 genotype. The cumulative distribution of age at onset is shown for RLS cases grouped by rs113851554 genotype dosage (0, 1, or 2 copies of the T allele). Age at onset analyses were performed in RLS cases from the All of Us cohort using linear regression adjusted for sex and the first ten principal components. The rs113851554 T allele was associated with a later age at onset (β = 1.54 years per allele, SE = 0.36; p = 2.03 × 10^-5^).

**Figure S6.**
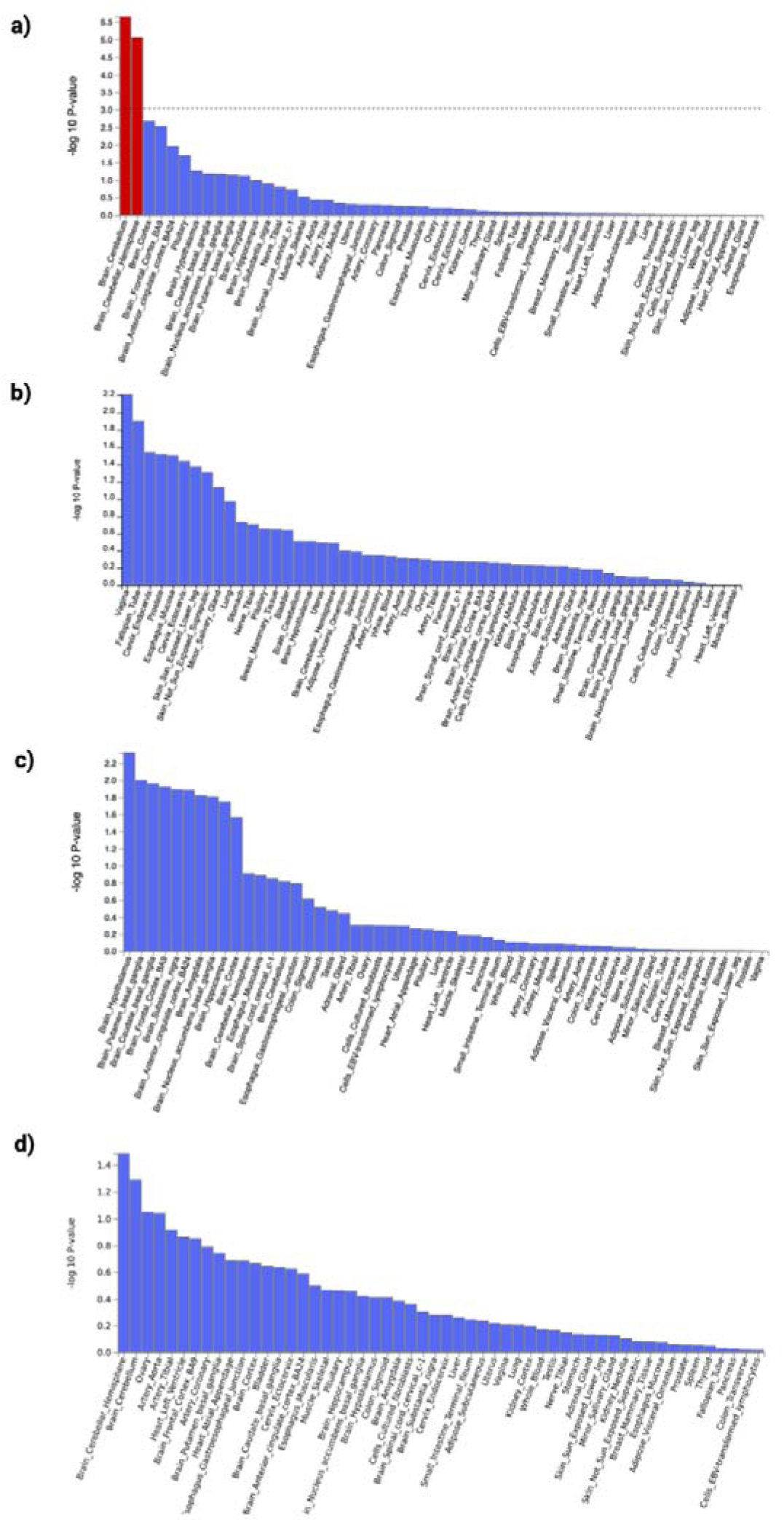
Tissue-specific expression enrichment of RLS-associated genes based on MAGMA analysis.

**Figure S7.**
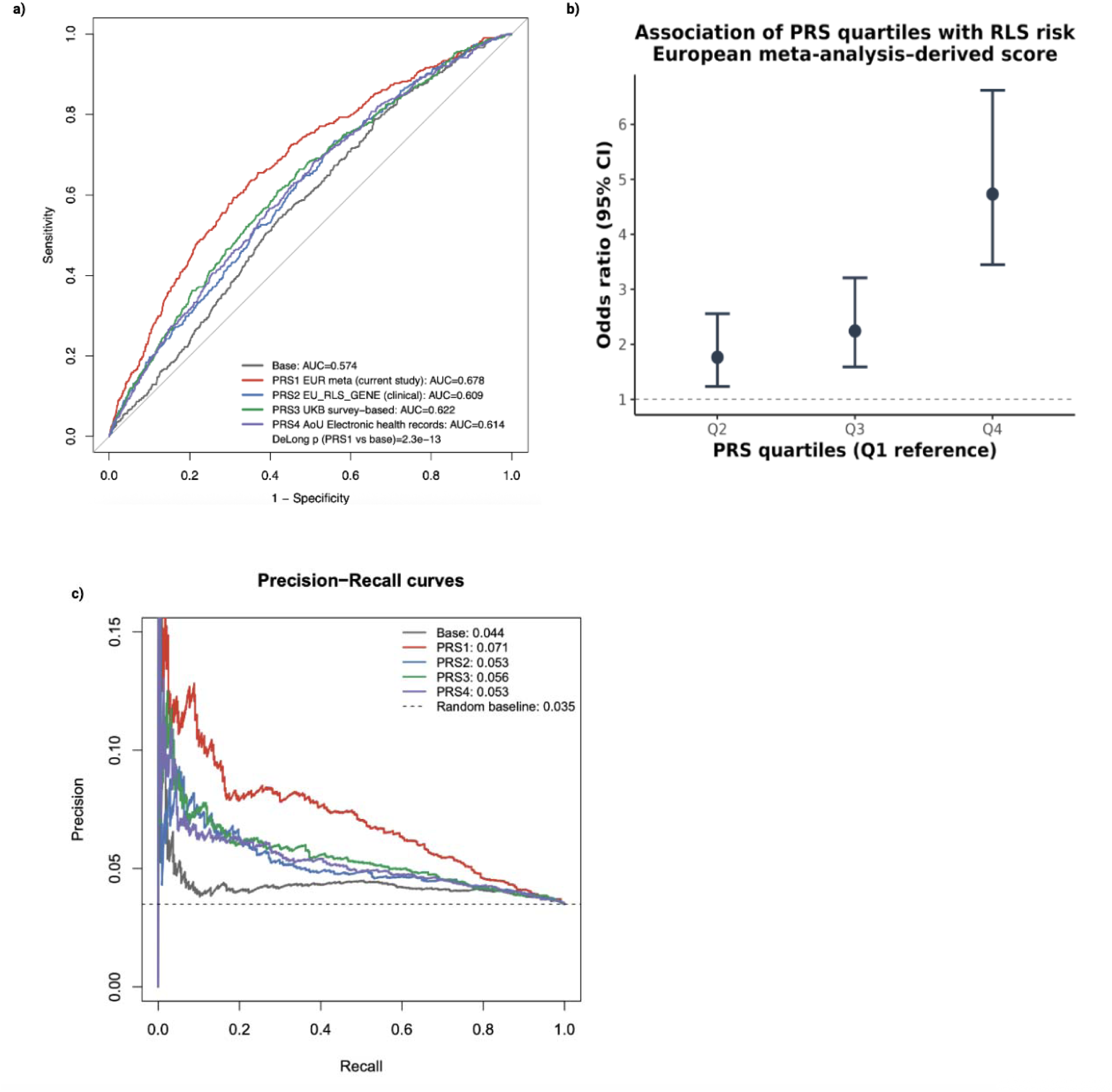
Performance of polygenic risk scores for RLS in an independent cohort. a) Receiver operating characteristic curves comparing base clinical model (age, sex, PCs) with PRS derived from four training GWAS datasets: European multi-cohort meta-analysis, EU-RLS-GENE (clinically ascertained), UK Biobank survey-based, and All of Us EHR-based. b) Association of European meta-analysis-derived PRS quartiles with RLS risk. Odds ratios were estimated using logistic regression adjusted for age, sex, and first ten PCs; Q1 used as reference. c) Precision-recall curves for the same models. The dashed horizontal line indicates the case prevalence in the target cohort, representing random classifier performance.

## References

1. Manconi, M. et al. Restless legs syndrome. *Nat Rev Dis Primers* **7**, 80 (2021).

2. Allen, R. P. et al. Restless legs syndrome/Willis-Ekbom disease diagnostic criteria: updated International Restless Legs Syndrome Study Group (IRLSSG) consensus criteria--history, rationale, description, and significance. Sleep Med 15, 860–873 (2014).

3. Winkelman, J. W. et al. Treatment of restless legs syndrome and periodic limb movement disorder: an American Academy of Sleep Medicine clinical practice guideline. J Clin Sleep Med 21, 137–152 (2025).

4. Xiong, L., et al. Family study of restless legs syndrome in Quebec, Canada: clinical characterization of 671 familial cases. Arch Neurol 67, 617–622 (2010).

5. Desautels, A. et al. Identification of a major susceptibility locus for restless legs syndrome on chromosome 12q. Am J Hum Genet 69, 1266–1270 (2001).

6. Pichler, I. et al. Linkage analysis identifies a novel locus for restless legs syndrome on chromosome 2q in a South Tyrolean population isolate. Am J Hum Genet 79, 716–723 (2006).

7. Edelson, J. L. et al. The genetic etiology of periodic limb movement in sleep. Sleep 46, (2023).

8. Akçimen, F. et al. Screening of novel restless legs syndrome-associated genes in French-Canadian families. Neurol Genet 4, e296 (2018).

9. Akçimen, F. et al. Genomic Analysis Identifies Risk Factors in Restless Legs Syndrome. Ann Neurol 96, 994–1005 (2024).

10. Akçimen, F. et al. Genetic and epidemiological characterization of restless legs syndrome in Québec. Sleep 43, (2020).

11. Tilch, E. et al. Identification of Restless Legs Syndrome Genes by Mutational Load Analysis. Ann Neurol 87, 184–193 (2020).

12. Didriksen, M. et al. Large genome-wide association study identifies three novel risk variants for restless legs syndrome. Commun Biol 3, 703 (2020).

13. Schormair, B. et al. Genome-wide meta-analyses of restless legs syndrome yield insights into genetic architecture, disease biology and risk prediction. Nat Genet 56, 1090–1099 (2024).

14. Schormair, B. et al. Identification of novel risk loci for restless legs syndrome in genome-wide association studies in individuals of European ancestry: a meta-analysis. Lancet Neurol 16, 898–907 (2017).

15. Fawale, M. B. et al. Restless legs syndrome: a rarity in the Nigerian pregnant population? Sleep Med 43, 47–53 (2018).

16. Burtscher, C. et al. Prevalence of restless legs syndrome in an urban population of eastern Africa (Tanzania). J. Neurol. Sci. 346, 121–127 (2014).

17. Castillo, P. R., Kaplan, J., Lin, S.-C., Fredrickson, P. A. & Mahowald, M. W. Prevalence of restless legs syndrome among native South Americans residing in coastal and mountainous areas. Mayo Clin Proc 81, 1345–1347 (2006).

18. Song, P. et al. The global and regional prevalence of restless legs syndrome among adults: A systematic review and modelling analysis. J. Glob. Health 14, 04113 (2024).

19. Verma, A. et al. Diversity and scale: Genetic architecture of 2068 traits in the VA Million Veteran Program. Science 385, eadj1182 (2024).

20. Winkelmann, J. et al. Genome-wide association study of restless legs syndrome identifies common variants in three genomic regions. Nat Genet 39, 1000–1006 (2007).

21. Tyrrell, J. et al. Genetic predictors of participation in optional components of UK Biobank. Nat Commun 12, 886 (2021).

22. Mägi, R. et al. Trans-ethnic meta-regression of genome-wide association studies accounting for ancestry increases power for discovery and improves fine-mapping resolution. Hum Mol Genet 26, 3639–3650 (2017).

23. Chang, C. C. et al. Second-generation PLINK: rising to the challenge of larger and richer datasets. Gigascience 4, 7 (2015).

24. Yuan, K. et al. Fine-mapping across diverse ancestries drives the discovery of putative causal variants underlying human complex traits and diseases. Nat Genet 56, 1841–1850 (2024).

25. Wallace, C. A more accurate method for colocalisation analysis allowing for multiple causal variants. PLoS Genet. 17, e1009440 (2021).

26. Wen, X., Pique-Regi, R. & Luca, F. Integrating molecular QTL data into genome-wide genetic association analysis: Probabilistic assessment of enrichment and colocalization. PLoS Genet. 13, e1006646 (2017).

27. Hormozdiari, F. et al. Colocalization of GWAS and eQTL signals detects target genes. Am. J. Hum. Genet. 99, 1245–1260 (2016).

28. Gusev, A. et al. Integrative approaches for large-scale transcriptome-wide association studies. Nat. Genet. 48, 245–252 (2016).

29. Zhu, Z. et al. Integration of summary data from GWAS and eQTL studies predicts complex trait gene targets. Nat. Genet. 48, 481–487 (2016).

30. Gamazon, E. R. et al. A gene-based association method for mapping traits using reference transcriptome data. Nat. Genet. 47, 1091–1098 (2015).

31. Liu, F. et al. Mitigating inconsistencies in GWAS follow-up analyses with LocusCompare2. Nat Genet (2025) doi:10.1038/s41588-025-02331-x.

32. All of Us Research Program Genomics Investigators. Genomic data in the All of Us Research Program. Nature 627, 340–346 (2024).

33. UK Biobank Whole-Genome Sequencing Consortium. Whole-genome sequencing of 490,640 UK Biobank participants. Nature 645, 692–701 (2025).

34. Raina, P. et al. Cohort Profile: The Canadian Longitudinal Study on Aging (CLSA). Int J Epidemiol 48, 1752–1753j (2019).

35. Awadalla, P. et al. Cohort profile of the CARTaGENE study: Quebec’s population-based biobank for public health and personalized genomics. Int J Epidemiol 42, 1285–1299 (2013).

36. Forgetta, V. et al. Cohort profile: genomic data for 26 622 individuals from the Canadian Longitudinal Study on Aging (CLSA). BMJ Open 12, e059021 (2022).

37. Vitale, D. et al. GenoTools: an open-source Python package for efficient genotype data quality control and analysis. G3 (Bethesda) 15, (2025).

38. Bulik-Sullivan, B. K. et al. LD Score regression distinguishes confounding from polygenicity in genome-wide association studies. Nat Genet 47, 291–295 (2015).

39. Karczewski, K. J. et al. Pan-UK Biobank genome-wide association analyses enhance discovery and resolution of ancestry-enriched effects. Nat Genet 57, 2408–2417 (2025).

40. Willer, C. J., Li, Y. & Abecasis, G. R. METAL: fast and efficient meta-analysis of genomewide association scans. Bioinformatics 26, 2190–2191 (2010).

41. Freedman, M. L. et al. Assessing the impact of population stratification on genetic association studies. Nat Genet 36, 388–393 (2004).

42. Lake, J. et al. Multi-ancestry meta-analysis and fine-mapping in Alzheimer’s disease. Mol Psychiatry 28, 3121–3132 (2023).

43. Kim, J. J. et al. Multi-ancestry genome-wide association meta-analysis of Parkinson’s disease. Nat Genet 56, 27–36 (2024).

44. Watanabe, K., Taskesen, E., van Bochoven, A. & Posthuma, D. Functional mapping and annotation of genetic associations with FUMA. Nat Commun 8, 1826 (2017).

45. Finan, C. et al. The druggable genome and support for target identification and validation in drug development. Sci Transl Med 9, (2017).

46. Mizuhara, E., Nakatani, T., Minaki, Y., Sakamoto, Y. & Ono, Y. Corl1, a novel neuronal lineage-specific transcriptional corepressor for the homeodomain transcription factor Lbx1. J Biol Chem 280, 3645–3655 (2005).

47. Sarayloo, F. et al. SKOR1 has a transcriptional regulatory role on genes involved in pathways related to restless legs syndrome. Eur J Hum Genet 28, 1520–1528 (2020).

48. Catoire, H. et al. A direct interaction between two Restless Legs Syndrome predisposing genes: MEIS1 and SKOR1. Sci Rep 8, 12173 (2018).

49. Nakatani, T., Minaki, Y., Kumai, M., Nitta, C. & Ono, Y. The c-Ski family member and transcriptional regulator Corl2/Skor2 promotes early differentiation of cerebellar Purkinje cells. Dev Biol 388, 68–80 (2014).

50. Rataj-Baniowska, M. et al. Retinoic Acid Receptor β Controls Development of Striatonigral Projection Neurons through FGF-Dependent and Meis1-Dependent Mechanisms. J Neurosci 35, 14467–14475 (2015).

51. Podleśny-Drabiniok, A. et al. Distinct retinoic acid receptor (RAR) isotypes control differentiation of embryonal carcinoma cells to dopaminergic or striatopallidal medium spiny neurons. Sci Rep 7, 13671 (2017).

52. Mapaga, J. N. et al. Prevalence of restless legs syndrome and associated factors in rural West African country. J. Neurosci. Rural Pract. 16, 224–230 (2025).

53. Fawale, M. B., Ismail, I. A., Mustapha, A. F., Komolafe, M. A. & Adedeji, T. A. Restless legs syndrome in a Nigerian elderly population. J. Clin. Sleep Med. 12, 965–972 (2016).

54. Liang, R. et al. Clinical features, polysomnography, and genetics association study of restless legs syndrome in clinic based Chinese patients: A multicenter observational study. Sleep Med 117, 123–130 (2024).

55. Kim, M.-K. et al. Association of restless legs syndrome variants in Korean patients with restless legs syndrome. Sleep 36, 1787–1791 (2013).

