## Supplementary Method for "Ancestry-specific and multi-ancestry genome-wide association studies of restless legs syndrome"

**SUPPLEMENTARY METHODS**

Study cohorts

The All of Us cohort (<https://allofus.nih.gov/>) comprises participants recruited from across the United States as part of a nationwide precision medicine initiative. We used data from the All of Us Research Program Controlled Tier Dataset v8. Participants were limited to those with available whole-genome sequencing and EHR data and genetically inferred ancestry corresponding to African (873 cases; 33,876 controls), Latin American (876 cases; 33,909 controls), and European (8,300 cases; 86,961 controls). RLS cases were identified using the SNOMED clinical terminology codes corresponding to RLS (SNOMED code: 32914008) from EHR condition records. Controls were defined as individuals without RLS-related diagnostic codes and without self-reported RLS symptoms. Individuals with missing age or sex information were excluded. Within the All of Us Research Program, all participants provided informed consent either in person or via an electronic consent (eConsent) platform, which includes primary consent and Health Insurance Portability and Accountability Act (HIPAA) authorization for the use of electronic health records in research. Data made available to researchers are de-identified prior to access and are analyzed within a controlled-access research environment under approved data use agreements. The study protocol was reviewed and approved by the Institutional Review Board (IRB) of the All of Us Research Program. The All of Us IRB operates in accordance with the regulations and guidance of the National Institutes of Health Office for Human Research Protections, ensuring consistent oversight and protection of participant rights and welfare^1^.

The VA Million Veteran Program ([https://www.mvp.va.gov](https://www.mvp.va.gov/)) contains EHRs and genotyping data of over 635,000 US veterans from diverse ancestral backgrounds. RLS case status was defined using a rule-based algorithm that relied solely on structured EHR data. Participants with two or more instances of International Classification of Diseases (ICD) codes corresponding to RLS (ICD-9: 333.94; ICD-10: G25.81) were classified as cases, whereas controls were defined as individuals with no recorded instances of these codes^2^. We utilized GWAS summary statistics from recently published analyses of African (1,303 cases; 119,437 controls), Latin American (873 cases; 33,876 controls), and European ancestry (1,148 cases; 57,993 controls) participants from the Million Veteran Program (MVP) dataset^3^. In the current study, only aggregate summary statistics derived from (MVP data were used, with no access to individual-level or identifiable participant data. Recruitment and enrollment for MVP were conducted within the Veterans Health Administration under an Institutional Review Board (IRB)-approved research protocol. MVP is a voluntary research program, and all participants provided informed consent along with Health Insurance Portability and Accountability Act (HIPAA) authorization prior to the use of their health information and biospecimens for research purposes. The program operates in accordance with VA research governance policies, including VHA Directives for the protection of human subjects, with oversight by the VA Central IRB to ensure ethical conduct, participant autonomy, and data protection throughout the research lifecycle. Further details on MVP recruitment, consent, transparency, and data governance are available in the publicly accessible Privacy Impact Assessment for the Patient Recruitment and Enrollment System for MVP (RNE) (https://department.va.gov/privacy/wp-content/uploads/sites/5/2025/08/FY25PatientRecruitmentandEnrollmentSystemforMVPRNEAssessingPIA.pdf).

The UK Biobank (<https://www.ukbiobank.ac.uk/>) is a large population-based resource that recruited approximately 500,000 participants aged 40-69 years from across the United Kingdom. Within this resource, a detailed sleep questionnaire was introduced to examine biological and environmental factors contributing to poor sleep and its consequences. The first set of this data was released to approved researchers in March 2025 and included nearly 180,000 UK Biobank participants. The sleep questionnaire included eight items to assess the five essential diagnostic criteria as defined by the IRLSSG^4^. Positive ascertainment for RLS was based on scoring criteria similar to that of the Cambridge-Hopkins diagnostic questionnaire for RLS, which has demonstrated a reasonable level of sensitivity and specificity for ascertainment of RLS as a patient completed questionnaire in population-based studies^5^. RLS was indicated if the participant responded positively to: (1) presence of the recurrent need to move the legs while sitting or lying down and/or recurrent uncomfortable feelings in legs while sitting or lying down; (2) the feelings present when resting; (3) feelings improve with movement; (4) feelings worse in the evening or at night; (5) feelings were not due to positional discomfort or leg cramps. We further limited cases to individuals reporting symptoms on several days per week, excluded those whose symptoms occurred only in the morning or persisted throughout the entire day, and required that symptoms were not usually relieved by changing leg position. A total of 7,317 RLS cases and 106,479 controls of European ancestry were included in final analyses. Participants of other ancestry groups were not analyzed due to limited sample sizes. Age was defined as age at recruitment (assessment center visit) and used as a covariate in all UK Biobank analyses. For the UK Biobank, all participants provided written informed consent, including consent for long-term follow-up through linkage to electronic health records and for the use of biological samples in health-related research. The UK Biobank study operates under an established Ethics and Governance Framework, with approval from the appropriate research ethics committees. Participant data and samples are held by UK Biobank as the legal custodian of the resource and are made available to approved researchers in anonymized form through controlled-access procedures under standard data access agreements. The study protocol and analytic use of UK Biobank data are conducted in accordance with the approved UK Biobank research protocol and governance framework.

The Canadian Longitudinal Study on Aging (CLSA, <https://www.clsa-elcv.ca/>) comprises individuals residing across Canada, whereas CARTaGENE (<https://cartagene.qc.ca/en>) includes participants residing in the province of Québec. Participants from both cohorts completed RLS-related questionnaire items. In CARTaGENE, case status was defined according to the essential diagnostic criteria of the IRLSSG, which evaluate the urge to move the legs, rest-induced symptoms, improvement with movement, evening or nighttime worsening, and exclusion of positional discomfort or cramps. In the CLSA, participants were asked a single screening question (“Do you have, or have you sometimes experienced, a recurrent need or urge to move your legs while sitting or lying down?”), capturing the first two IRLSSG criteria.^6^. The RLS-related diagnostic questions and corresponding criteria are summarized in Table S10. GWAS summary statistics from these cohorts were utilized in this study, as previously described in Akçimen et al. (2024), where we generated the initial GWAS results^7^, which included 921 RLS cases and 1,307 controls from CARTaGENE and 4,980 cases and 15,990 controls from the CLSA.

For the CARTaGENE cohort, participants consented to the long-term use of their data and biological samples for future health and genomic research. All collected data and samples are coded to ensure confidentiality. CARTaGENE is a publicly funded research infrastructure managed by the Université de Montréal, which is responsible for governance, including data and sample security. Participants aged 40 to 69 years residing in selected metropolitan areas of Quebec were recruited to obtain a representative population sample and received detailed study information, including consent materials, before participation. The participant electronically signed the consent form. Access to CARTaGENE data and samples is granted only to researchers whose projects have been approved by appropriate research ethics committees (https://cartagene.qc.ca/files/documents/consent/brochure_en_0505.pdf). For the CLSA, participants provided signed informed consent prior to participation, in accordance with the CLSA Privacy Policy (https://www.clsa-elcv.ca/wp-content/uploads/2023/06/privacy_policy.pdf). The study includes both a tracking component, involving telephone interviews, and a comprehensive component, involving in-home interviews, data collection site visits, physical assessments, and optional blood and urine samples.

GWAS of participation in the UK Biobank RLS diagnostic questionnaire

To evaluate potential participation bias in the questionnaire-derived phenotype, we conducted a genome-wide association study of participation in the UK Biobank RLS diagnostic survey. The analysis was performed across unrelated European ancestry UK Biobank European participants who were eligible for the questionnaire. Individuals who completed the RLS diagnostic survey were coded as participants (n = 155,705), and those who did not complete the survey were coded as non-participants (n = 264,439).

Genetic association testing was performed using PLINK v2.0 using a logistic regression, adjusting for age, sex, genotyping array, and the first ten principal components. Sample- and variant-level quality control procedures were identical to those applied in the primary UK Biobank GWAS described in Methods. This analysis therefore evaluates genetic influences on survey participation independent of disease status. Genome-wide significant loci identified in the participation GWAS were subsequently compared with loci identified in the RLS case-control GWAS.

External GWAS summary statistics

Genotyping of MVP samples was performed using the custom ThermoFisher Axiom MVP 1.0 array. Samples with a call rate below 98.5% were removed^8^. Imputation was conducted using a hybrid reference panel composed of the African Genome Resources panel^9^ and the 1000 Genomes Project Phase 3 Version 5 reference panel^10^. Post-imputation variant level quality control was applied to exclude variants with a squared correlation coefficient (r^2^) < 0.3, minor allele count below 20, call rate below 97.5% for common variants. In addition, variants showing greater than 10% deviation from their expected allele frequency based on 1000 Genomes reference data were removed^3^. Prior to meta-analysis, we excluded variants with effect allele frequency < 0.01 or > 0.99 from the MVP summary statistics to remove low- or high-frequency variants and avoid unstable effect estimates.

Genotyping for the CLSA cohort was performed using the Affymetrix Axiom 2.0 array (794,409 markers; 29,970 participants)^11^. Samples with genotype missingness greater than 5% and discordance between reported and genetically inferred sex were excluded. Pairwise relatedness and genetic ancestry were assessed using KING^12^, and unrelated participants of European ancestry were retained for downstream analyses by removing one individual from each related pair (kinship coefficient >0.125, which corresponds to the expected kinship for second-degree relationships in KING^13^. Variant-level quality control was conducted in PLINK v2^14^, removing multiallelic and non-autosomal variants, as well as variants with call rate < 99%, minor allele frequency < 0.01, evidence of differential missingness between cases and controls (P < 1.0 × 10^-4^), or significant deviation from Hardy-Weinberg equilibrium in controls (P < 1.0 × 10^-10^)^7^.

Genotyping for the CARTaGENE cohort was carried out using the Illumina Infinium Global Screening Array (GSAMD-24v1-0_20011747_A1; 700,078 markers; 2,228 participants). Cohort characteristics and quality control procedures have been detailed previously by Awadella et al^15^. The same sample and variant-level filtering pipeline applied to the CLSA dataset was used for CARTaGENE. Samples with >5% missingness or sex discordance were removed. Relatedness and ancestry were evaluated with KING^12^, and unrelated European ancestry individuals were included after removing one individual from each related pair (KINSHIP > 0.125, corresponding to the expected kinship for second-degree relatives)^13^. Variants failing call rate (<99%), minor allele frequency (<0.01), differential missingness (P < 1.0 × 10^-4^), or Hardy-Weinberg equilibrium thresholds (P < 1.0 × 10^-10^) were excluded. Genetic ancestry for CARTaGENE and CLSA was inferred by projecting study participants onto principal components derived from the 1000 Genomes Project reference panel.

Imputation for CARTaGENE and CLSA had been previously performed using the same pipeline on the TOPMed Imputation Server with Eagle v2.4 phasing and the TOPMed reference panel (hg38)^16–18^. Post-imputation, variants with minor allele frequency <0.5% and squared correlation coefficient (r^2^) < 0.5 were removed^7^^,^^12^.

For the CLSA and CARTaGENE cohorts, we used our previously published GWAS summary statistics of European ancestry^7^. For the MVP, ancestry-specific summary statistics were obtained from Verma et al. (2023) via the GWAS Catalog (GCST90477503, GCST90475828, GCST90475829)^3,7^. For cohorts analyzed using previously published summary statistics, covariate adjustment followed the analytical frameworks of the original studies, which included age, sex, and ancestry principal components selected to account for population structure (CARTaGENE: sex, age, PC1, PC2, PC4, PC7, PC10; Canadian Longitudinal Study on Aging: sex, age, PC2, PC6). GWASs in the MVP cohort were performed using the SAIGE mixed model framework^19^ to account for case-control imbalance and sample relatedness, with adjustment for age, sex, and 10 PCs^3^. Variants with effect allele frequency < 0.01 or > 0.99 were filtered out from the MVP GWAS summary statistics.

Polygenic risk score analysis

A separate cohort was constructed for polygenic risk score analyses using European ancestry participants from the All of Us who were not included in the GWAS analyses. Genotyping data were generated using the Illumina Global Diversity Array (1,825,277 markers/GDA-8 v1.0)^20^. The array cohort consisted of 441 RLS cases and 12,172 controls. Related individuals were removed using a kinship threshold of 0.0084^12^. Standard genotype-level quality control was applied using PLINK v2^14^, excluding variants with minor allele frequency <0.01, genotype missingness >1%, or deviation from Hardy-Weinberg equilibrium P < 1×10^-6^. Imputation was performed using the TOPMed Imputation Server with Eagle phasing and the TOPMed reference panel^16^^,^^17^. Variants with a squared correlation coefficient (r^2^) < 0.8 were removed before PRS construction.

Polygenic risk scores were generated using PRS-CS^21^, a Bayesian regression framework with continuous shrinkage priors, which models local linkage disequilibrium using the 1000 Genomes European reference panel. Four independent training GWAS summary statistics were evaluated: (i) the European multi-cohort meta-analysis from the current study (All of Us WGS, MVP, UK Biobank, CLSA, CARTaGENE); (ii) the clinically ascertained EU-RLS-GENE GWAS reported by Schormair et al., 2024^22^; (iii) a survey-based UK Biobank GWAS; and (iv) an electronic health record–based All of Us GWAS from the current study. For each training dataset, posterior SNP effect sizes estimated by PRS-CS were applied separately to the target genotype data using PLINK v2 score function^14,22^. Scores were aligned to the effect allele, standardized to mean 0 and standard deviation 1 within the target cohort, and evaluated under identical covariate adjustment. Association between PRS and RLS status was tested using logistic regression adjusted for age, sex, and the first ten PCs. Discriminative performance was quantified by area under the ROC curve (AUC). Improvement beyond the base model was assessed using likelihood-ratio tests and ΔAUC. To evaluate risk stratification, PRS deciles were constructed within the target cohort and odds ratios comparing the highest versus lowest decile were estimated from covariate-adjusted models. Statistical comparisons of ROC curves were performed using DeLong’s test^23^.

**REFERENCES**

1	Doerr M, Grayson S, Moore S, Suver C, Wilbanks J, Wagner J. Implementing a universal informed consent process for the Research Program. *Pac Symp Biocomput* 2019; **24**: 427–38.

2	Denny JC, Bastarache L, Ritchie MD, *et al.* Systematic comparison of phenome-wide association study of electronic medical record data and genome-wide association study data. *Nat Biotechnol* 2013; **31**: 1102–10.

3	Verma A, Huffman JE, Rodriguez A, *et al.* Diversity and scale: Genetic architecture of 2068 traits in the VA Million Veteran Program. *Science* 2024; **385**: eadj1182.

4	Allen RP, Picchietti DL, Garcia-Borreguero D, *et al.* Restless legs syndrome/Willis-Ekbom disease diagnostic criteria: updated International Restless Legs Syndrome Study Group (IRLSSG) consensus criteria--history, rationale, description, and significance. *Sleep Med* 2014; **15**: 860–73.

5	Allen RP, Burchell BJ, MacDonald B, Hening WA, Earley CJ. Validation of the self-completed Cambridge-Hopkins questionnaire (CH-RLSq) for ascertainment of restless legs syndrome (RLS) in a population survey. *Sleep Med* 2009; **10**: 1097–100.

6	Allen RP, Picchietti D, Hening WA, *et al.* Restless legs syndrome: diagnostic criteria, special considerations, and epidemiology. A report from the restless legs syndrome diagnosis and epidemiology workshop at the National Institutes of Health. *Sleep Med* 2003; **4**: 101–19.

7	Akçimen F, Chia R, Saez-Atienzar S, *et al.* Genomic Analysis Identifies Risk Factors in Restless Legs Syndrome. *Ann Neurol* 2024; **96**: 994–1005.

8	Hunter-Zinck H, Shi Y, Li M, *et al.* Genotyping Array Design and Data Quality Control in the Million Veteran Program. *Am J Hum Genet* 2020; **106**: 535–48.

9	Sengupta D, Botha G, Meintjes A, *et al.* Performance and accuracy evaluation of reference panels for genotype imputation in sub-Saharan African populations. *Cell Genom* 2023; **3**: 100332.

10	1000 Genomes Project Consortium, Auton A, Brooks LD, *et al.* A global reference for human genetic variation. *Nature* 2015; **526**: 68–74.

11	Forgetta V, Li R, Darmond-Zwaig C, *et al.* Cohort profile: genomic data for 26 622 individuals from the Canadian Longitudinal Study on Aging (CLSA). *BMJ Open* 2022; **12**: e059021.

12	Manichaikul A, Mychaleckyj JC, Rich SS, Daly K, Sale M, Chen W-M. Robust relationship inference in genome-wide association studies. *Bioinformatics* 2010; **26**: 2867–73.

13	Zhang Q-X, Jayasinghe D, Zhang Z, Lee SH, Xu H-M, Chen G-B. Precise estimation of in-depth relatedness in biobank-scale datasets using deepKin. *Cell Rep Methods* 2025; **5**: 101053.

14	Chang CC, Chow CC, Tellier LC, Vattikuti S, Purcell SM, Lee JJ. Second-generation PLINK: rising to the challenge of larger and richer datasets. *Gigascience* 2015; **4**: 7.

15	Awadalla P, Boileau C, Payette Y, *et al.* Cohort profile of the CARTaGENE study: Quebec’s population-based biobank for public health and personalized genomics. *Int J Epidemiol* 2013; **42**: 1285–99.

16	Taliun D, Harris DN, Kessler MD, *et al.* Sequencing of 53,831 diverse genomes from the NHLBI TOPMed Program. *Nature* 2021; **590**: 290–9.

17	Fuchsberger C, Abecasis GR, Hinds DA. minimac2: faster genotype imputation. *Bioinformatics* 2015; **31**: 782–4.

18	Das S, Forer L, Schönherr S, *et al.* Next-generation genotype imputation service and methods. *Nat Genet* 2016; **48**: 1284–7.

19	Zhou W, Nielsen JB, Fritsche LG, *et al.* Efficiently controlling for case-control imbalance and sample relatedness in large-scale genetic association studies. *Nat Genet* 2018; **50**: 1335–41.

20	All of Us Research Program Genomics Investigators. Genomic data in the All of Us Research Program. *Nature* 2024; **627**: 340–6.

21	Ge T, Chen C-Y, Ni Y, Feng Y-CA, Smoller JW. Polygenic prediction via Bayesian regression and continuous shrinkage priors. *Nat Commun* 2019; **10**: 1776.

22	Schormair B, Zhao C, Bell S, *et al.* Genome-wide meta-analyses of restless legs syndrome yield insights into genetic architecture, disease biology and risk prediction. *Nat Genet* 2024; **56**: 1090–9.

23	DeLong ER, DeLong DM, Clarke-Pearson DL. Comparing the areas under two or more correlated receiver operating characteristic curves: a nonparametric approach. *Biometrics* 1988; **44**: 837–45.
