## Supplementary figures and images for "Ancestry-specific and multi-ancestry genome-wide association studies of restless legs syndrome"

### Supplementary Figure 1

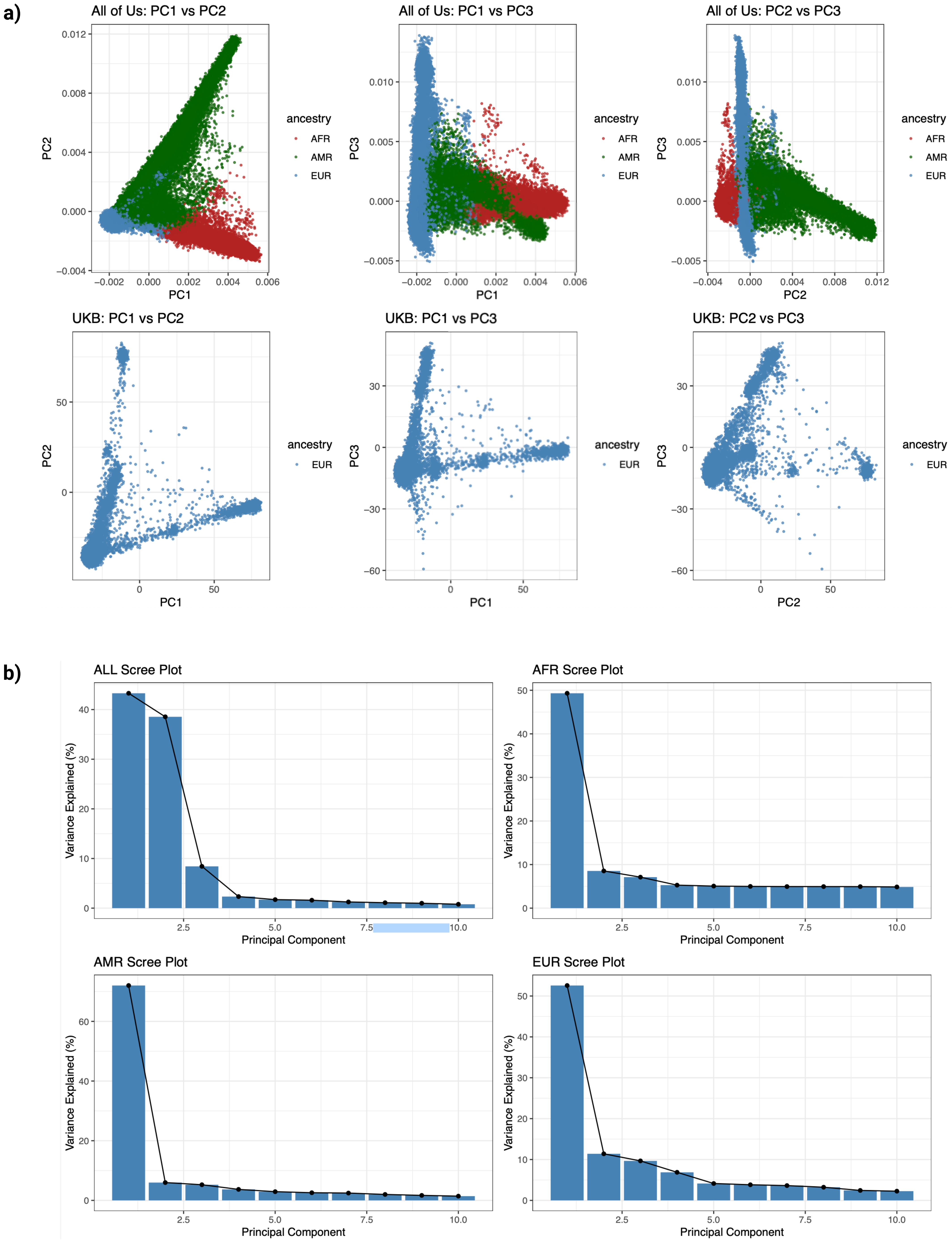

### Supplementary Figure 2

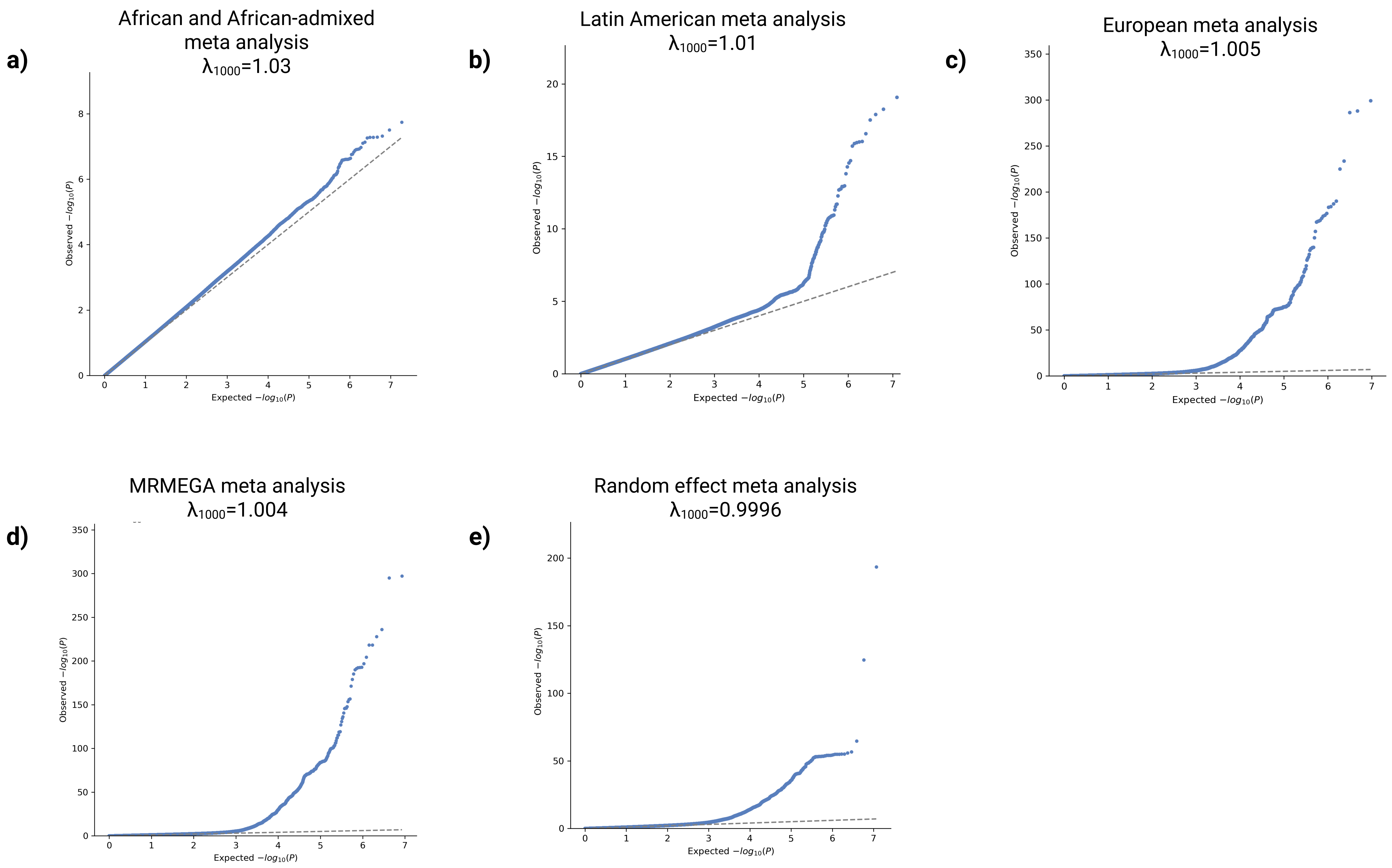

### Supplementary Figure 3

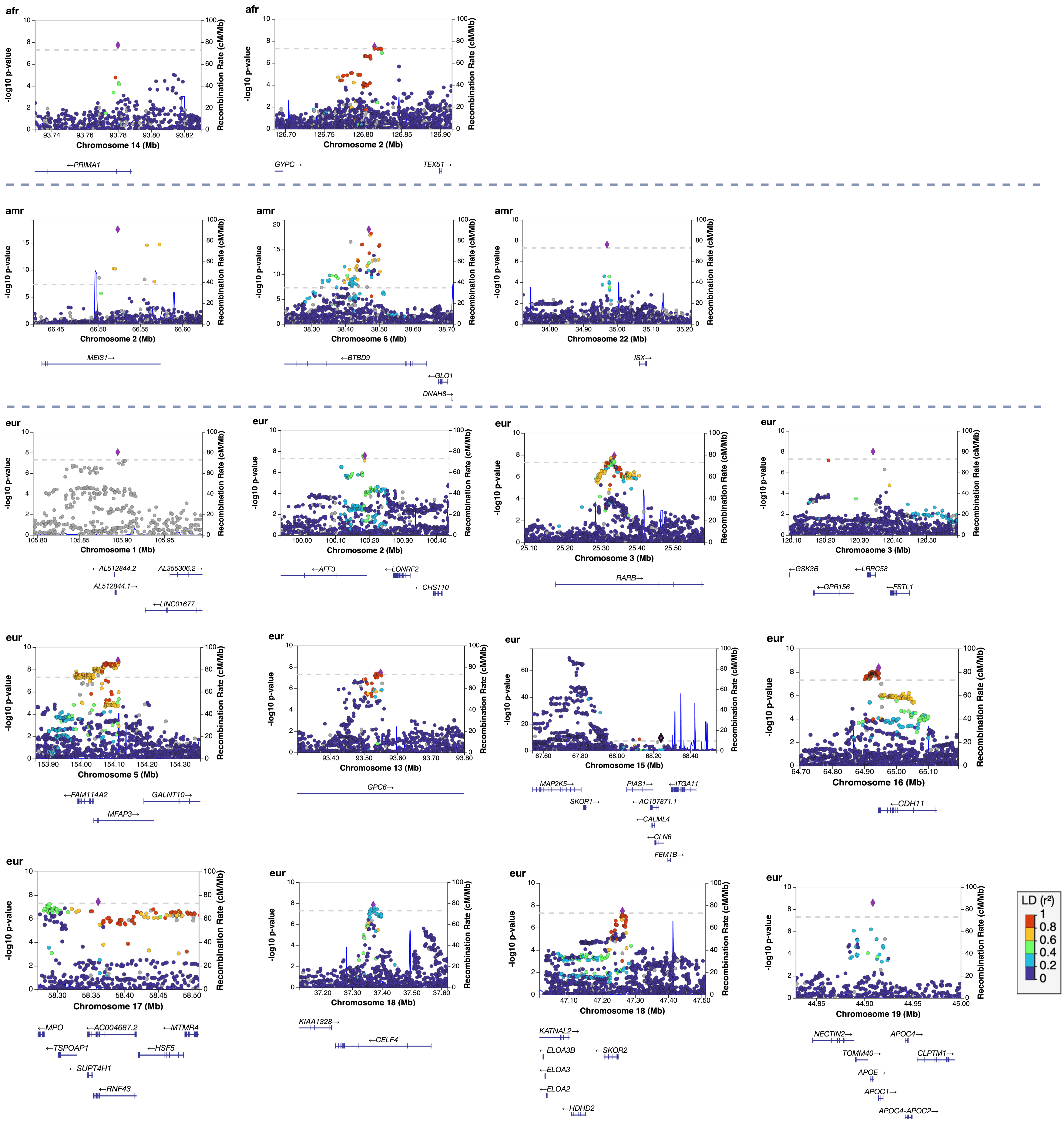

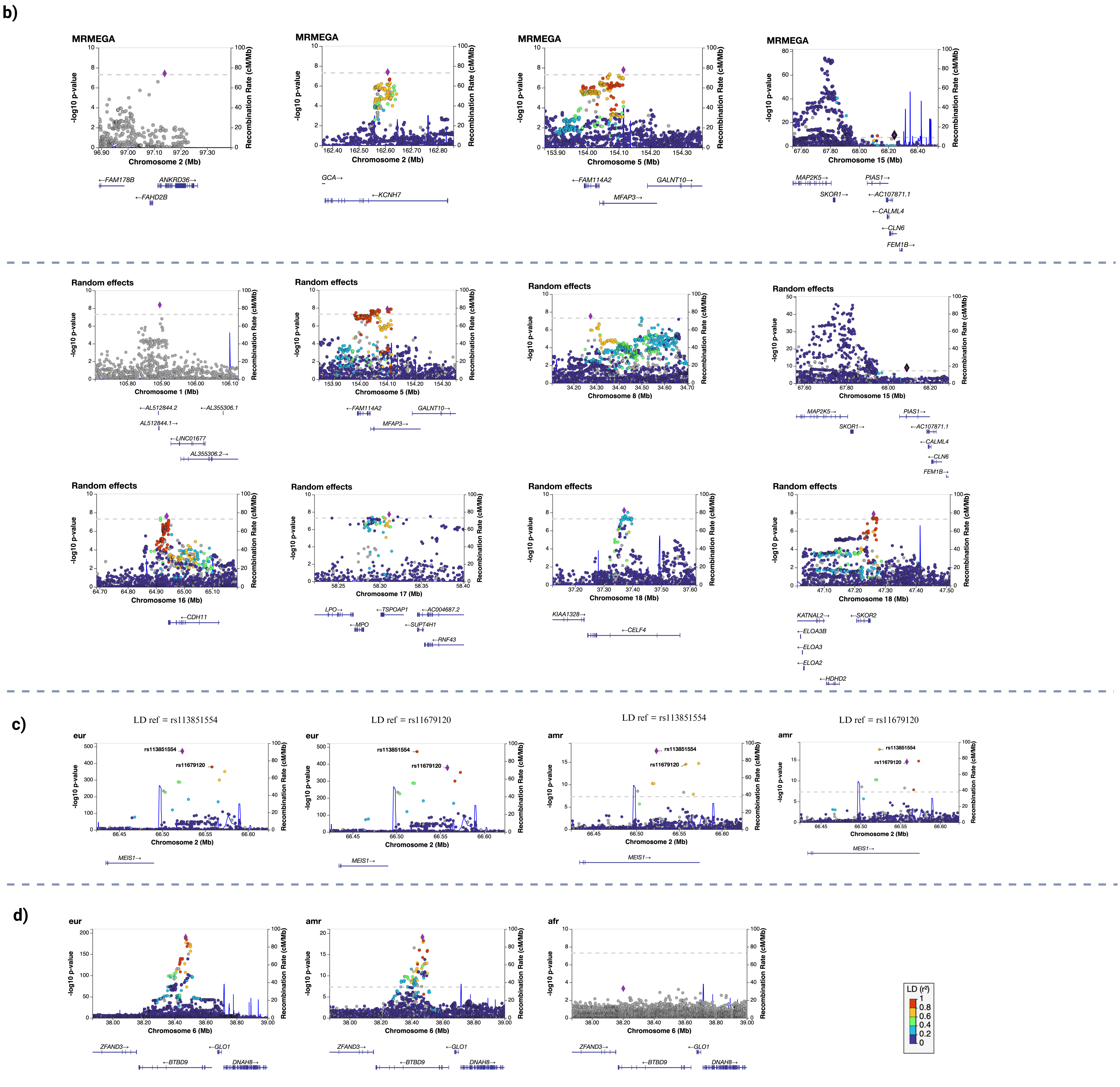

### Supplementary Figure 4

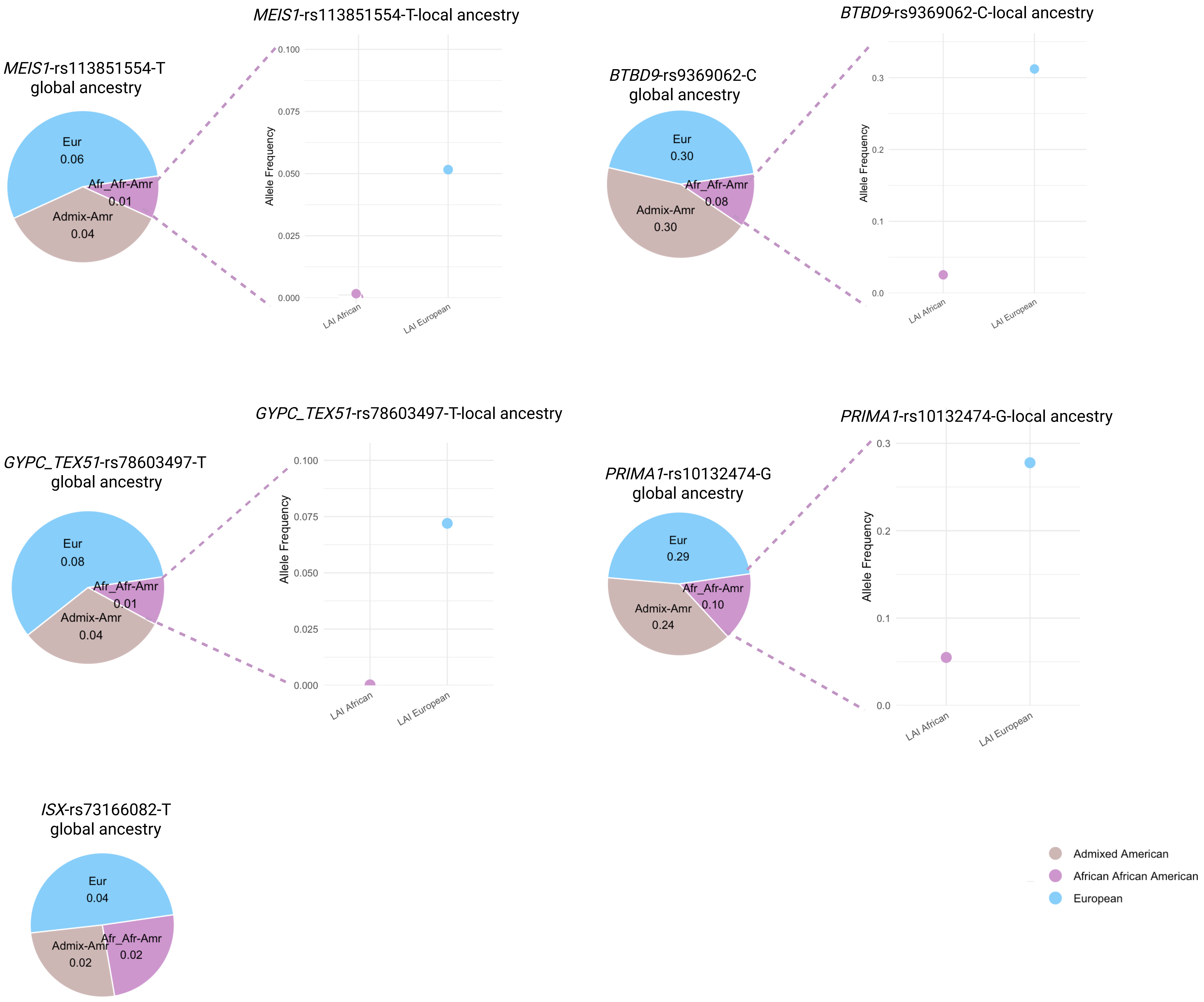

### Supplementary Figure 5

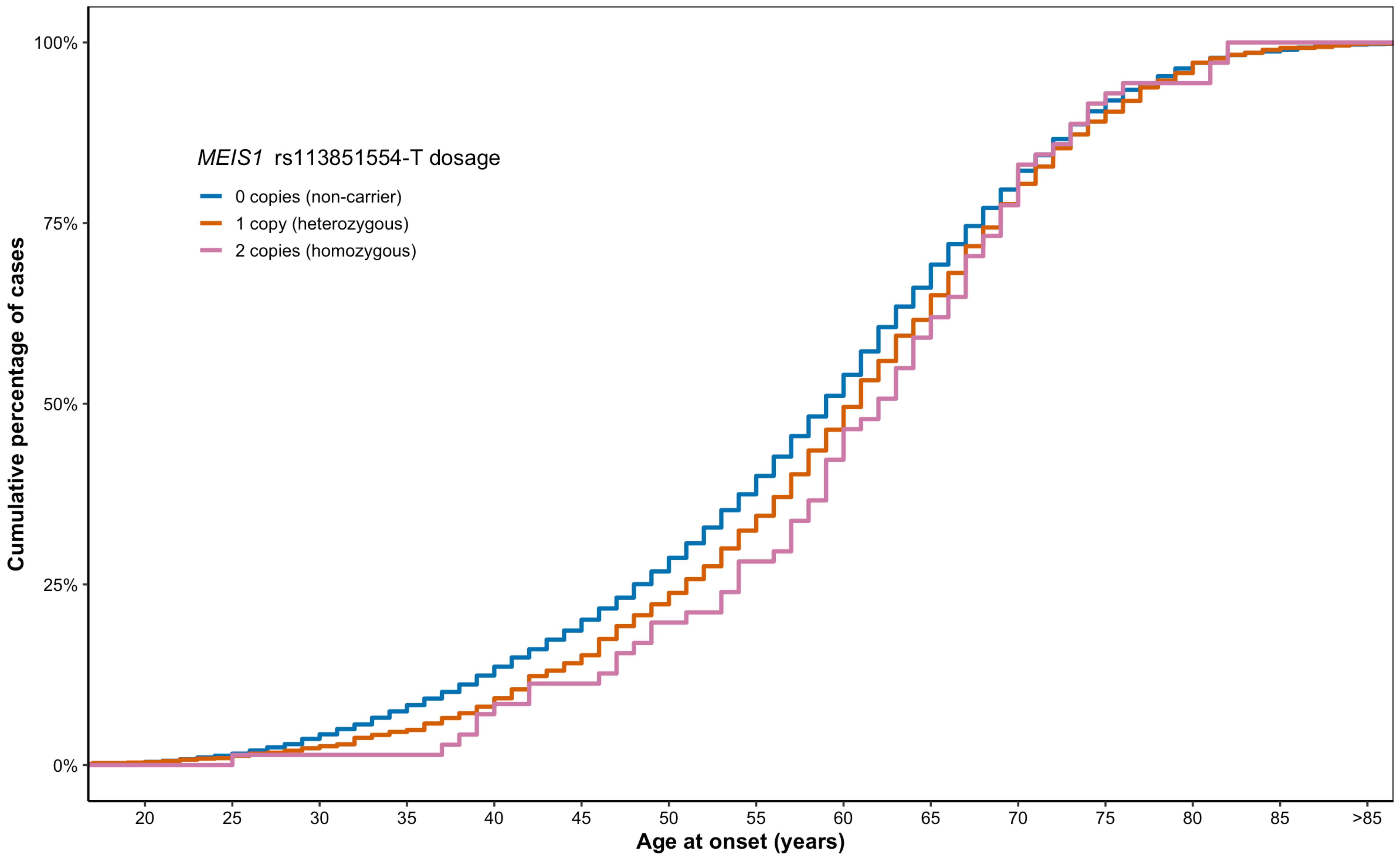

### Supplementary Figure 6

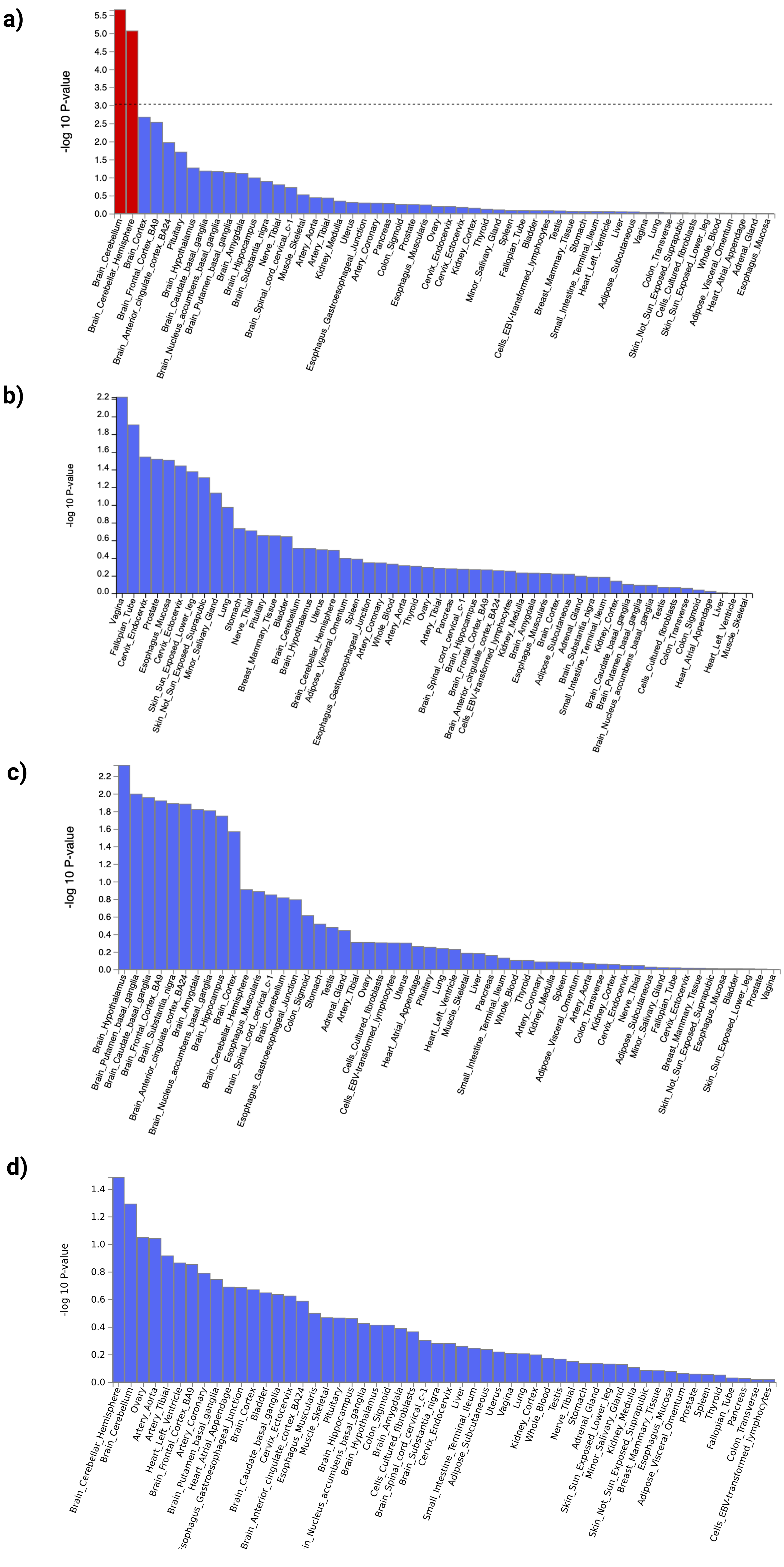

### Supplementary Figure 7

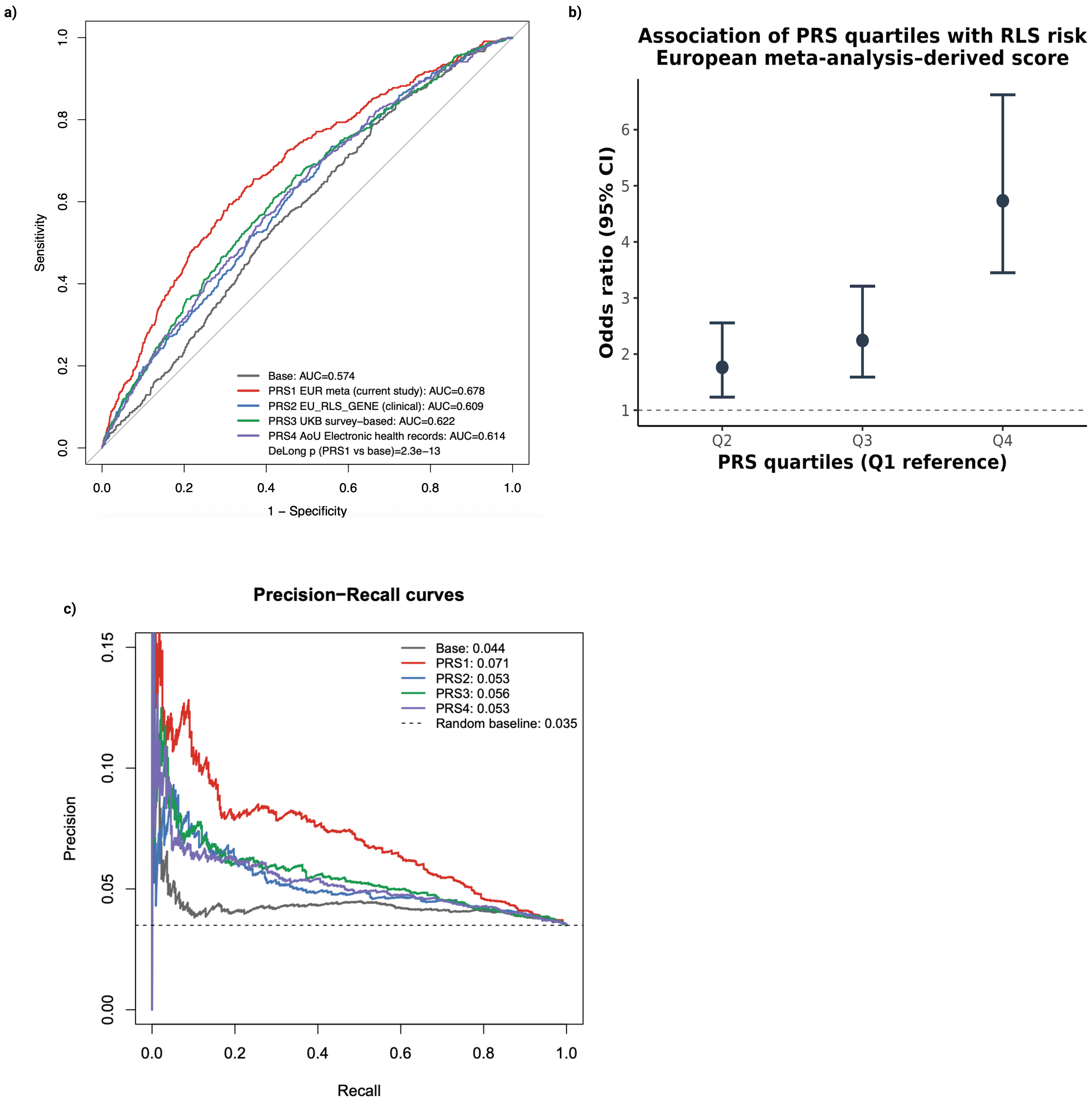
